# Intracalvariosseous injection bypasses the blood-brain barrier as a novel drug delivery approach for pre-clinical and clinical trials in stroke

**DOI:** 10.1101/2024.03.03.24303548

**Authors:** Wenqian Liu, Nanxing Wang, Mo Yang, Meiyang Zhang, Xiangrong Liu, Kaibin Shi, Weiming Liu, Yunwei Ou, Liping Liu, Zhonghua Yang, Yanfang Liu, Mengyuan Zhou, Xiaoling Liao, Hui Qu, Fu-Dong Shi, Yuesong Pan, Chaoyu Wang, Xuezheng Feng, Xingquan Zhao, Mingjun Zhang, Yongjun Wang, Yilong Wang

**Affiliations:** Department of Neurology, Beijing Tiantan Hospital, Capital Medical University, Beijing, China; Chinese Institute for Brain Research, Beijing, China; National Center for Neurological Disorders, Beijing, China; Advanced Innovation Center for Human Brain Protection, Capital Medical University, Beijing, China; China National Clinical Research Center for Neurological Diseases, Beijing, China; Beijing Laboratory of Oral Health, Capital Medical University, Beijing, China; Department of Neurosurgery, Beijing Tiantan Hospital, Capital Medical University, Beijing, China; Laboratory for Clinical Medicine, Capital Medical University, Beijing, China; Department of Biomedical Engineering, School of Medicine, Tsinghua University, Beijing, China

**Keywords:** Blood-brain barrier, Microchannels, Intracalvariosseous injection, Drug delivery, Stroke

## Abstract

Central nervous system (CNS) accessibility constitutes a major hurdle for drug development to treat neurological diseases. Existing drug delivery methods rely integrity of the blood-brain barrier (BBB) for CNS penetration. Here we showed that the microchannels between the skull marrow and the dura mater could be harnessed for drug delivery by intracalvariosseous (ICO) injection. Drugs administered via ICO injection were found to reach cranial bone marrow-dura-perivascular space, and the injection procedure did not cause osteomyelitis or BBB damage. To validate this approach, we examined the efficacy of two neuroprotective agents, NA-1 and Y-3, via ICO injection in rat model of stroke and found that ICO injection increased drug accumulation in the brain compared to intravenous injection, reduced infarct area and alleviated neurological deficits. We subsequently conducted a clinical trial to assess the safety of ICO in acute ischemic stroke patients (ClinicalTrials.gov identifier NCT05849805), showing that ICO injection was feasible and safe in humans and its therapeutic effects may be observed. Collectively, our study identifies that the microchannels between the skull bone marrow and the dura mater act as a new channel for CNS drug delivery to achieve high intracranial drug exposure in a short period of time. The safety of ICO injection makes it a promising route of drug administration for CNS diseases.

## Introduction

Treatments of central nervous system (CNS) diseases often require CNS accessibility of the drugs. However, the existence of the blood-brain barrier (BBB) allows only a small fraction of hydrophobic small molecules to enter the brain. In contrast, most macromolecular drugs and biological agents such as peptides, proteins, and monoclonal antibodies cannot pass through BBB^1^. To address this challenge, recent studies have investigated non-invasive methods including focused ultrasound combined with microbubble technology to open the BBB, nasal inhalation administration and nano dosage forms to increase drug bioavailability, etc^2–7^. However, safety issues relating to these techniques remain. For example, although focused ultrasound can enhance drug entry into the brain, it carries the risk of partially or fully opening of the BBB for a period of 6-24 hours^8^. This extended time window poses a significant threat to the alter CNS microenvironment given the pivotal role of the BBB in maintaining the appropriate conditions for neuronal activity^9^. In addition, opening the BBB may introduce immune cells or microorganisms into the brain tissues. Therefore, it is imperative to explore alternative drug delivery approaches that enhance drug access without compromising the integrity of the BBB.

In 2018, Herisson et al.^10^ made a significant discovery regarding the existence of microchannels connecting the skull bone marrow and the brain. Recent studies on the microchannels have further supported this finding. For instance, Pulou et al.^11^ reported that a fluorescent tracer injected into the cisterna magna of mice migrated through the perivascular space of the dural blood vessels and entered the skull bone marrow via these skull microchannels and so indicating that cerebrospinal fluid (CSF) can enter the skull bone marrow. Mazzitell et al.^12^ discovered that after spinal cord injury, CSF signals promote myelopoiesis and the entry of myeloid cells into the meninges. These findings revealed a mechanism of communication between the CNS and cranial bone marrow through CSF, which regulates the immune response of the CNS. Kang et al.^13^ have demonstrated the potential of utilizing microchannels between the skull bone marrow and the brain surface through an intracalvariosseous (ICO) method for drug delivery in rodents. It is worth noting that the ICO method they tested requires thinning the skull and mounting the device within the initial 24 hours, makes it more suitable for treating chronic neurological diseases than acute conditions such as strokes and brain injury.

In this study, we aimed to optimize the application of ICO for the pharmacological treatment of acute ischemic stroke and maximize clinical benefit by reducing the overall procedure time. To this end, we evaluated an improved injection method of ICO by using skull single-hole injection, which has the potential to reduce the extent of skull damage and shorten the procedure time to roughly 40 minutes, which broadened the application of ICO to acute CNS diseases such as acute ischemic stroke. Actually, malignant middle cerebral artery infarction (mMCAI) accounts for 10% of ischemic stroke cases and has a mortality rate as high as 40-60%^14,15^ and morbidity so resulting in a significant disease burden; further thrombolysis and endovascular therapy are not recommended for as treatment^16,17^. Decompressive craniectomy (DC) is currently the primary therapeutic intervention for mMCAI^18^, although DC does not reduce the infarction volume and may lead to infection and direct tissue injury^19^. Neuroprotection would be an attractive approach to manage mMCAI but no FDA approved therapies are available, which may be due to inadequate drug delivery through the BBB. So, we conducted preclinical experiments and subsequent clinical trial to prove feasibility, safety, and efficacy studies of ICO for the first time in a stroke animal model and also in mMCAI patients in a Phase I clinical trial called “Y-3 Injection Through Skull Bone Marrow in the Treatment of Acute Malignant Middle Cerebral Artery Infarction” (SOLUTION) (ClinicalTrials.gov identifier NCT05849805). Additionally, we hypothesized that after ICO injection, the drug might bypass the BBB through microchannels between the skull and dura mater to enter the brain parenchyma without affecting BBB permeability, which has been verified by fluorescence tracing the drug delivery pathway after ICO injection.

## Results

### Intracalvariosseous (ICO) injection can deliver dye molecules to the brain parenchyma

To examine whether the skull bone marrow-dura-glymphatic pathway can be utilized to transport drugs, we injected 5 μl of 10% Evans blue (EB) solution into mice via either tail vein or bone marrow through skull sigle-hole injection (Fig. 1a). Blue staining of the mouse limbs and tails were observed at 10 minutes, 30 minutes, and 60 minutes after injection through the tail vein, but not in the brain tissue. Blue staining of mouse tail veins could be seen at 30 and 60 minutes after the ICO injections, and blue staining of mouse brain was noted at 10-60 minutes after injections (Fig. 1b-c). It’s intriguing that when mice were injected with the same volume of EB solution, the concentration of EB was higher in those who received ICO injection compared to intravenous (IV) injection, in all tested brain areas such as skull and dura mater, cerebellum, cortex, striatum, and hippocampus, except for plasma. The EB content peaked at 1 h post ICO injection, and then gradually decreased. Furthermore, we observed that the fluorescence intensity of EB in the brain tissue near the site of occipital injection was higher than that in distant brain region (Extended Data Fig. 1a-b), we also noted that EB staining was mainly concentrated in the subcortical area.

**Fig. 1.**
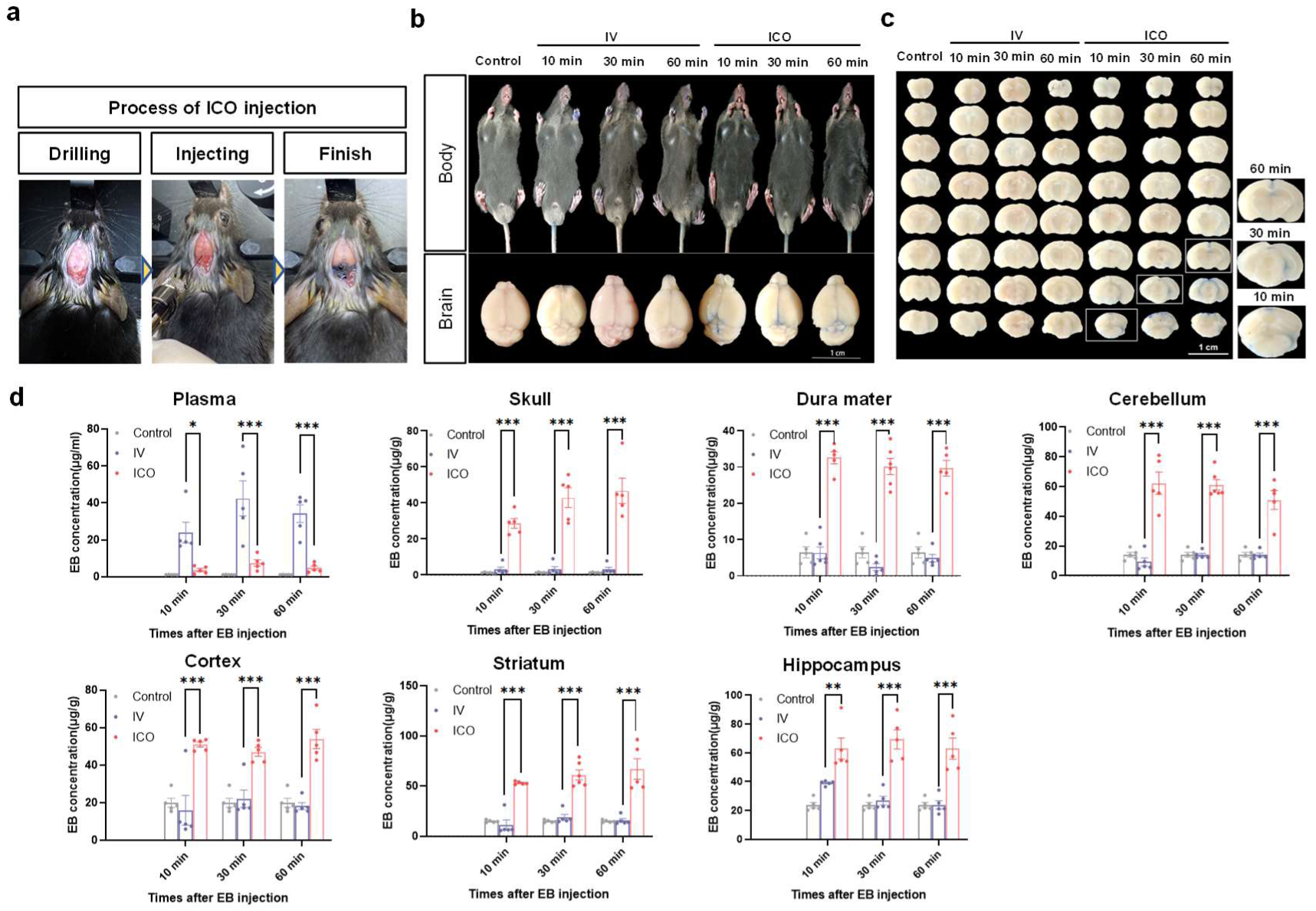
Feasibility of ICO injection. **a.** Operation diagrams of ICO injections: (1) opening hole, (2) drug injection, (3) Completion of the injection. **b-c.** Blue staining of mouse limbs and brain tissues at different time points 10 min, 30 min, and 60 min after EB injection by ICO or IV, the Control group did not inject EB, scale=1 mm. **d.** EB content in different tissues (plasma, skull, dura mater, cerebellum, cortex, corpus striatum, and hippocampi) of mice at 10 min, 30 min, and 60 min after EB injections by ICO or IV. Data are mean ± SEM. Two-way ANOVA (Tukey’s multiple comparisons test). *p<0.05, **p<0.01, ***p<0.001.

To assess single hole ICO injection side effects, we examined skin and subcutaneous tissues surrounding the injection sites and counted neutrophils in skull bone marrow and peripheral blood (Extended Data Fig. 2a-b). No redness, swelling or exudation were observed at the injection sites 24 hours after ICO injection in sham or pMCAO mice. Flow cytometry revealed that there were no significant changes in skull bone marrow and peripheral blood neutrophil cell numbers 24 hours after ICO injection (Extended Data Fig. 2c), suggested that the ICO injection did not cause skull osteomyelitis. The increase in peripheral blood neutrophils and the decrease in skull bone marrow neutrophils after the pMCAO model were likely caused by active neutrophil mobilization and migration from remote bone marrow site such as femur and tibia, or from the calvarium^10^.The integrity of BBB was tested by injecting FITC-dextran of different molecular weights (4k Da, 40k Da, 150k Da) via the tail vein 1 hour after the ICO injection of EB (Extended Data Fig. 2d). The results of 4k Da, 40k Da, and 150k Da (Extended Data Fig. 2e) FITC-dextran all supported that ICO did not induce BBB leakage into perivascular space.

### ICO injection bypassed BBB to reach the brain parenchyma

To delineate the delivery pathway of ICO injection to the brain, it is necessary to comprehensively examine the skull, dura mater and brain tissue. We employed a Cy3 labeled polypeptide PSD95 inhibitor NA-1, C105H188N42O30, molecular weight: 2518.88. After ICO injection of Cy3-NA1, the skull-dura mater-brain complex was obtained 1 hour later and subjected to tissue transparency to alleviate imaging issues caused by the presence of calcified skull (Fig. 2a-b). It was found that Cy3-NA1 was present in the skull, microchannels between the skull and dura mater, dura mater, and brain parenchyma, with a higher concentration in the skull marrow than brain parenchyma. Additionally, 488-Lyve1 and 647-CD31 were used to label meningeal lymphatic vessels and blood vessels in the cleared tissue, respectively (Fig. 2c). Cy3-NA1 was found to be located around brain parenchyma and the blood vessels in the skull marrow cavity and microchannels, co-stained with Lyve1 (Fig. 2d), suggested that the fluorescence-labeled drug has entered the brain parenchyma through the microchannels in the skull marrow cavity and so bypassing the BBB. We found that ICO injection of drugs did not rely on the blood pathway, but rather bypassed the BBB via the perivascular space, which is consistent with CSF entering dura through spaces around the blood vessels^11^.

**Fig. 2.**
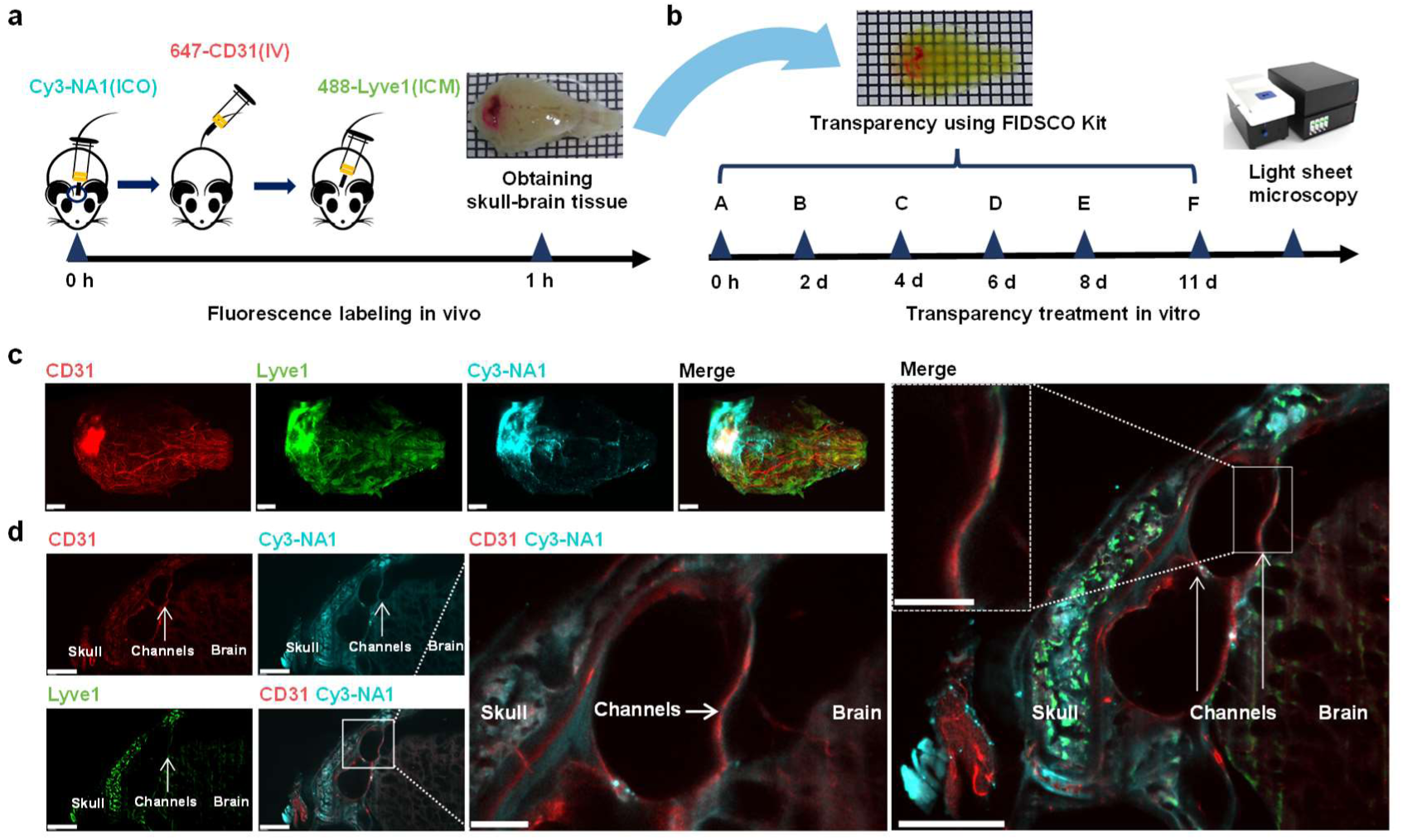
Cy3 labeled NA-1 injected by ICO enters brain parenchyma through the microchannels between skull and dura mater. **a.** Process of injected Cy3-NA1 by ICO, 647-CD31 by intravenous and 488-Lyve1 by intracisternal magna. **b.** Skull-dura-brain integrated tissue processing timeline. **c.** Overall scan results of lymphatic vessels, blood vessels and Cy3-NA1, scale=2 mm. **d.** Cy3-NA1 entered brain tissue through microchannels (white arrows) between the skull and brain parenchyma after ICO injection, scale=1 mm, inset=300 μm.

### Feasibility, safety, and efficacy of NA-1/Y-3 with ICO injection in stroke animal models

The feasibility, safety, and efficacy of ICO injection were evaluated in the rat pMCAO model of stroke, and PSD95 inhibitors were selected as the therapeutic drug for stroke. By inhibiting PSD95, the production of PSD95-NMDR-nNOS complex can be disrupted, leading to a reduction in neurotoxic damage^20^. NA-1(a polypeptide, C105H188N42O30, molecular weight: 2518.88) and Y-3 (a chemical compound, C24H27Cl2NO4, molecular weight: 463.13) are two different types PSD95 inhibitors. Preclinical studies have demonstrated that administering NA1/Y-3 during the acute stage of stroke can minimize temporary and permanent damage^21–24^. After 24 hours of administration, the brain tissue was stained with TTC to label the area of brain infarction. Compared with the IV group, the ICO group had a lower infarct area and mNSS score when the dose of NA-1 or Y-3 were 3.9 mg/kg or 0.1 mg/kg respectively, however the doses of IV group were 7.8 mg/kg or 4 mg/kg for NA-1 or Y-3 respectively (Fig. 3a-b). Treated rats were followed up for 7 days (Fig. 3c). Although there were no differences in survival curve (Fig. 3d) or weight changes (Fig. 3e) between the ICO and IV groups, the NA-1/Y-3 ICO group showed better neurological function score on the first and third days (Fig. 3f) and a longer time to drop in the Rotarod test on day three (Fig. 3g). We observed that ICO injection of NA-1 and Y-3 resulted in decreases in brain infarct area of 40% and 37% respectively compared to IV injection (Fig. 3h). These data suggested that the ICO injection of the neuroprotective agents was feasible, safe, and efficient as a treatment of stroke. The concentration of Y-3 in the tissues was tested by high performance liquid chromatography (HPLC) after IV/ICO injection. We found that the plasma Y-3 concentration in the ICO group was lower than that in the IV group at 1 hour to 4 hours after injection in the stroke model (Extended Data Fig. 3a). Moreover, the Y-3 distribution, as indicated by the tissue-to-plasma partition coefficient (*K*p), demonstrated a declining trend in the skull, dura mater, and CSF within the ICO group (Extended Data Fig. 3b), whereas an increasing trend was observed in the cerebellum, cortex, striatum, hippocampus, and deep cervical lymph nodes (Extended Data Fig. 3c). Additionally, we used FITC labeled NA-1 and Y-3 to observe the amount of drug injected by ICO, and found that ICO resulted in an increased number of FITC molecules and neurons with intracellular FITC compared to IV group (Extended Data Fig. 4a-b). Additionally, we assessed the liver and kidney functions in different injection groups and found no differences between ICO group and IV group (Extended Data Fig. 5a-d).

**Fig. 3.**
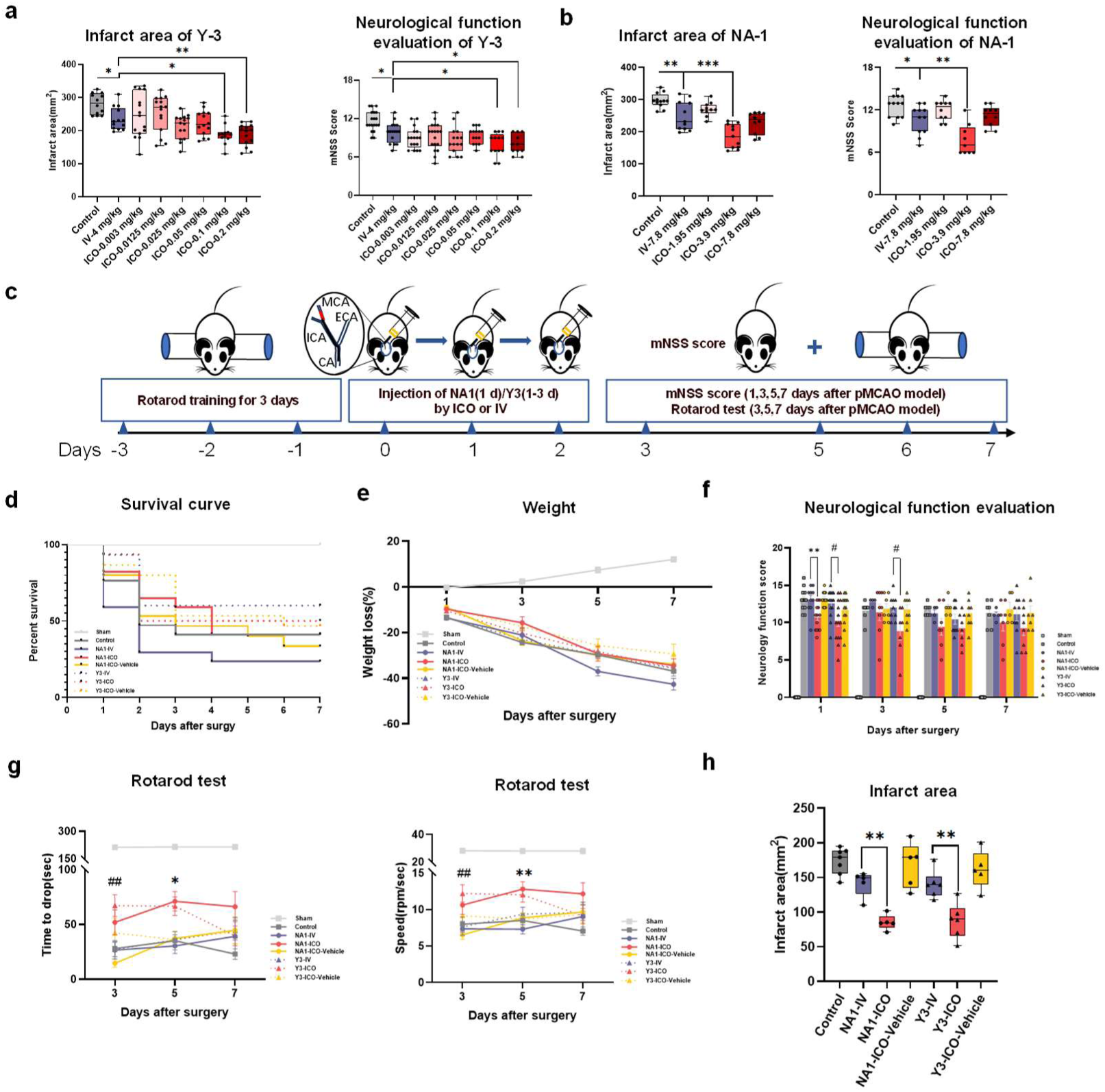
The efficacy of the neuroprotective agents NA-1/Y-3 in the pMCAO model. **a.** 24h efficacy of different doses of Y-3 injected IV or ICO in the pMCAO model, infarct area(left) and mNSS score (right) (n=10-15). **b.** 24h efficacy of different doses of NA-1 injected IV or ICO in the pMCAO model, infarct area(left) and mNSS score (right)(n=10-15). **c.** 7-day timeline of pMCAO model treated with NA-1/Y-3 via ICO injection. **d-h.** General conditions and neurological functions of pMCAO model improved after the ICO injection of Y-3(*n=14-16*). (d) Survival curve, (e) Changes of body weight, (f) mNSS score, (g) Rotarod test residence time and speed. (h) Brain infarct area. Data are mean ± SEM. (a, b, h) One-way ANOVA (Tukey’s multiple comparisons test). *p<0.05, **p<0.01. d. Log-rank (Mantel Cox) test. (e-g) Two-way ANOVA (Tukey’s multiple comparisons test). NA-1: IV vs ICO *p<0.05, **p<0.01; Y-3: IV vs ICO #p<0.05, ##p<0.01.

Since the PSD95 inhibitors can uncouple PSD95 and NMDR, and inhibit the expression of nNOS after strokes, we examined nNOS in the brain tissue of the pMCAO model 24 hours after injections of NA-1/Y-3 (Extended Data Fig. 4c). Semi-quantitative analysis of the fluorescence intensity demonstrated that the expression of nNOS in the brain tissue decreased after NA-1/Y-3 injection through ICO (Extended Data Fig. 4d). After strokes, neurons gradually experience programmed cell death due to ischemia and hypoxia. Capase3 and NeuN immunofluorescence staining were used to co-label apoptotic neurons in the pMCAO model (Extended Data Fig. 4e). The data showed apoptotic cells and apoptotic neurons decreased after the Y-3 injection into the skull bone marrow at day 7 (Extended Data Fig. 4f). In brief, ICO injection of the PSD95 inhibitors appeared to reduce the expression of nNOS and subsequent neuronal apoptosis after stroke.

### Clinical Trial: Demographic and baseline characteristics of patients

Between April 14, 2023 and November 6, 2023, a total of 161 stroke patients were screened, of whom 20 patients who were not suitable or responsive to reperfusion therapy (12.4%) were enrolled in the trial (Extended Data Fig. 6). All patients completed a 14-day follow-up period, except for three patients who succumbed to the stroke or other diseases within this timeframe. Among the screened patients, 72 were excluded for not meeting the inclusion criteria (Supplemental Table 3), and 69 were not included due to the presence of the exclusion criteria. Among all the ICO patients, 8 successfully underwent three sessions of ICO Y-3 administration. Two patients received two treatment sessions each; one due to a DC following neurosurgical assessment, and the other due to a fatal posterior circulation infarction. The demographic characteristics of the two patient groups were mostly similar (Extended Table 1). The median age of all participants was 58 years (IOR, 56 to 65), and 6 (30%) of 20 patients were women. The median age of the conventionally treated group was non-significantly younger (median age 57 vs 63, p=0.17). Six (30%) patients received intravenous thrombolysis with alteplase, and one patient in the ICO group received endovascular therapy with poor outcome prior to the enrollment. The mean time from onset to randomization was 13.8 hours (SD 3.7) while this was shorter in the conventional treatment (median time 12 vs 15, p=0.06). The mean volume of hypoperfusion areas was 268.0 ml (SD 73.8). Conventional treatment group had smaller hypoperfusion volume than the ICO group (median 236 vs 291, p=0.11), implicated potentially worse outcomes of the ICO group. The mean infarcted core volume was 121.2 ml (SD 58.8) and the median NIHSS score was 20 (IQR, 18 to 22), which were similar between the two groups. The spontaneous reperfusion conditions of the two groups were the same.

**Table 1.**
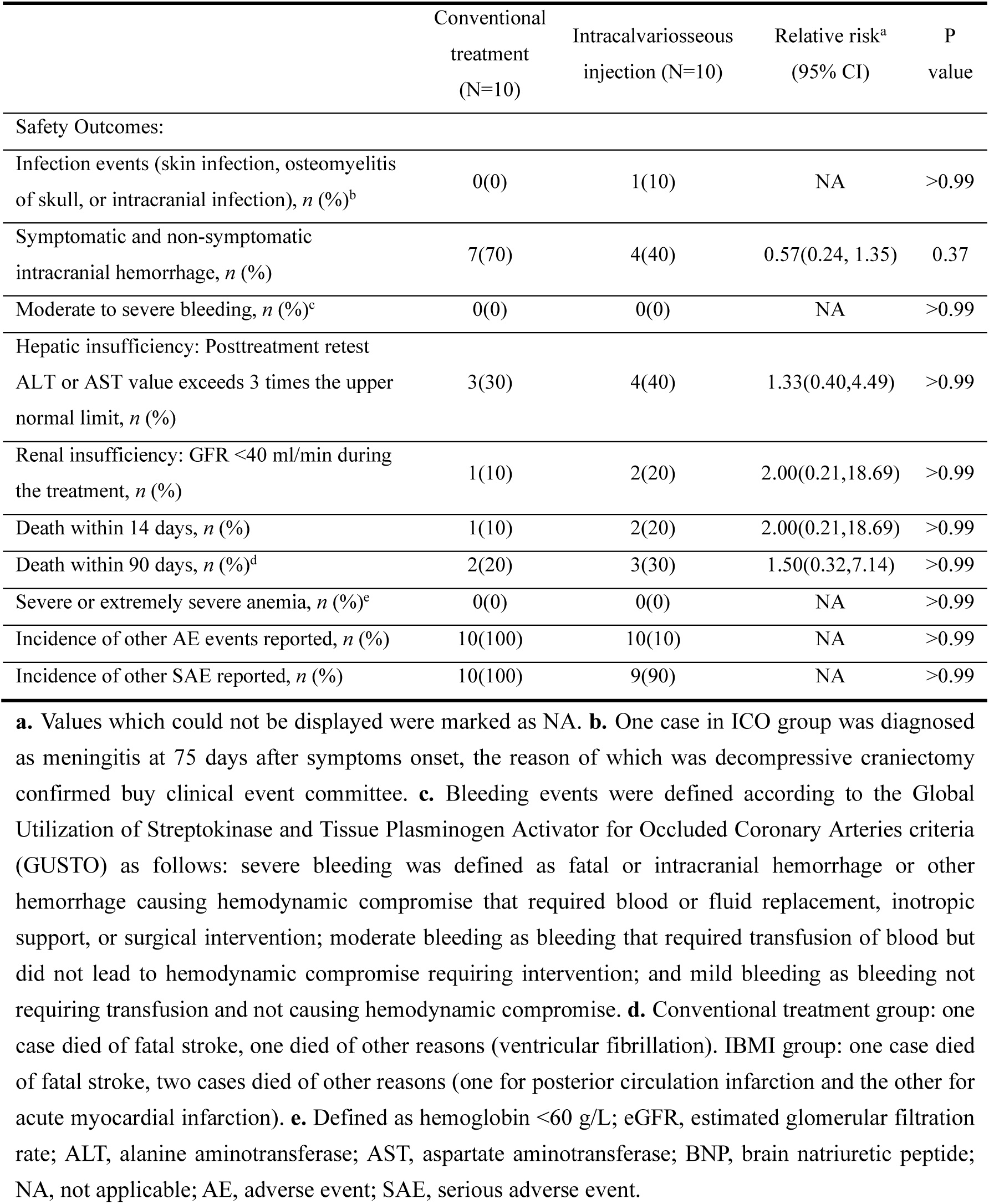
Safety outcomes.

### Primary Outcomes: safety of ICO in stroke patients

Of the 10 ICO group patients, ICO injections were successfully performed in all cases. The primary steps of operations are shown in fig. 4a. Drilling appearance are shown in fig. 4b-d. A cumulative of 12 skull drilling episodes (the skull of one patient was drilled on the first two days of the administration, and the skull of another patient was drilled twice in one operation) and 28 administrations of Y-3 were achieved in the ICO group (Extended Table 2). Neither skull inner plate penetration nor interruptions of the procedure due to drug leakage or patients’ non-cooperation were observed (Extended Table 2). The median of injection volume was 1.2ml (IQR, 1.0 to 1.3). The procedure was performed at the bedside inside the ICU but required an isolated aseptic operating space. The median duration of drilling was 32 minutes (IQR, 22 to 48). The median time from the disinfection to the drilling was 19 minutes (IQR, 14 to 33) and the median time from drilling to injection was 3 minutes (IQR, 2 to 10). Operations without drilling took a median of 22 minutes (IQR, 20 t0 28) while from start to injection took 13 minutes (IQR, 11 to 15) on average. The duration of the operation was related to the operator’s level of proficiency and events encountered during the procedure (such as skull hole obstruction). In addition, the median of door to injection time (DIT) was 4.7 hours (IQR, 4.2 to 6.6), including time for thrombolysis assessment, endovascular therapy, DC, and hospitalization procedures. In summary, skull outer plate drilling and ICO could be performed at the bedside and was completed in 30 minutes and all processes from diagnosis to completion of treatment could be completed within 5 hours upon hospital arrival.

**Fig. 4.**
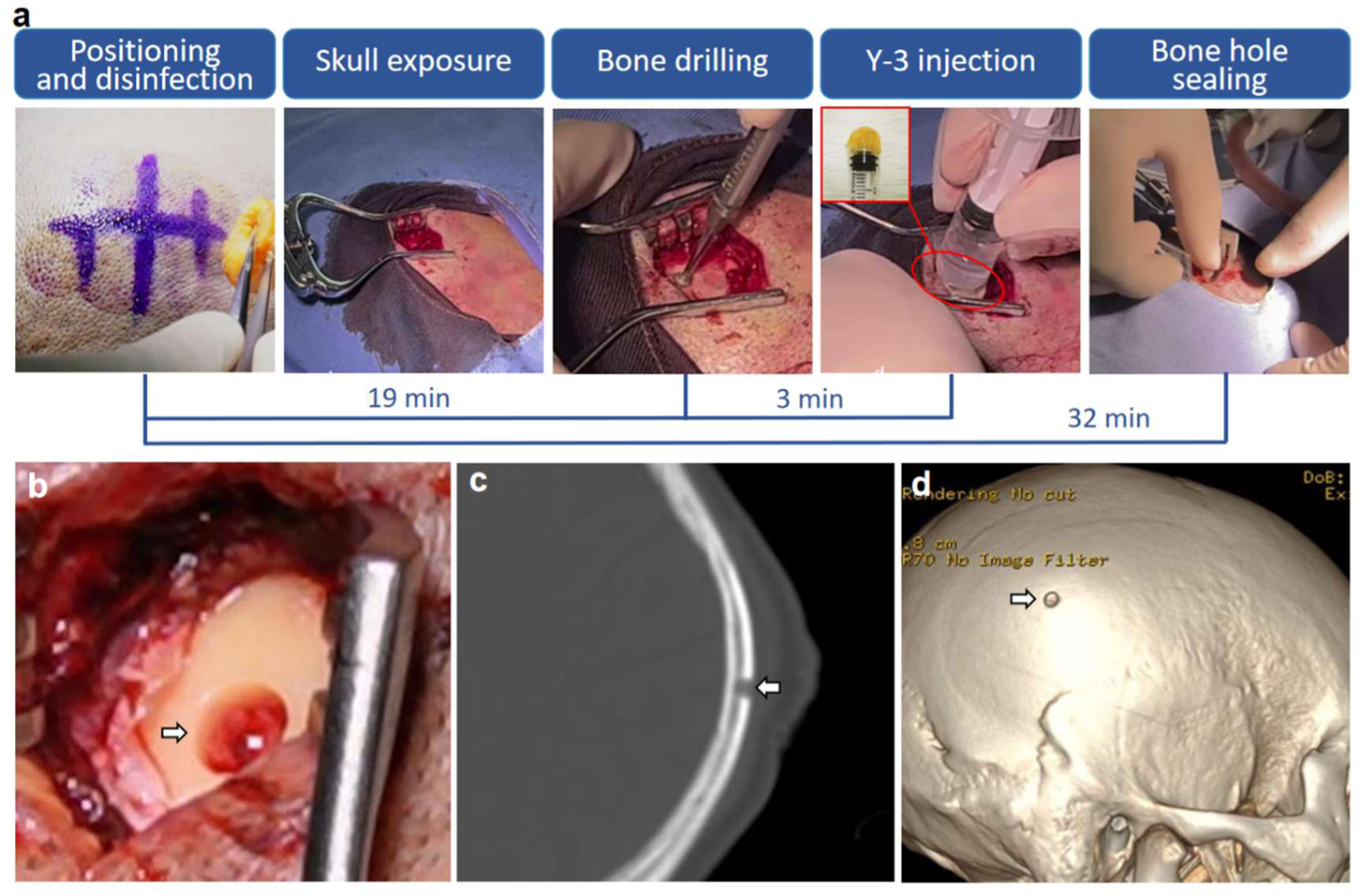
ICO injection of Y-3 in a stroke patient. **a.** The procedure and duration of the ICO protocol. Head cone of the syringe was covered by three layers of foam dressing. **b.** Drilling hole (white arrow) on surgical field of the skull. **c.** Appearance of the drilling hole (white arrow) shown in computed tomography (CT) bone window. The drilling only penetrated the outer plate of the skull without affecting the inner plate. d. Appearance of drilling hole (white arrow) on three-dimensional reconstruction of skull with CT thin section.

**Table 2.**
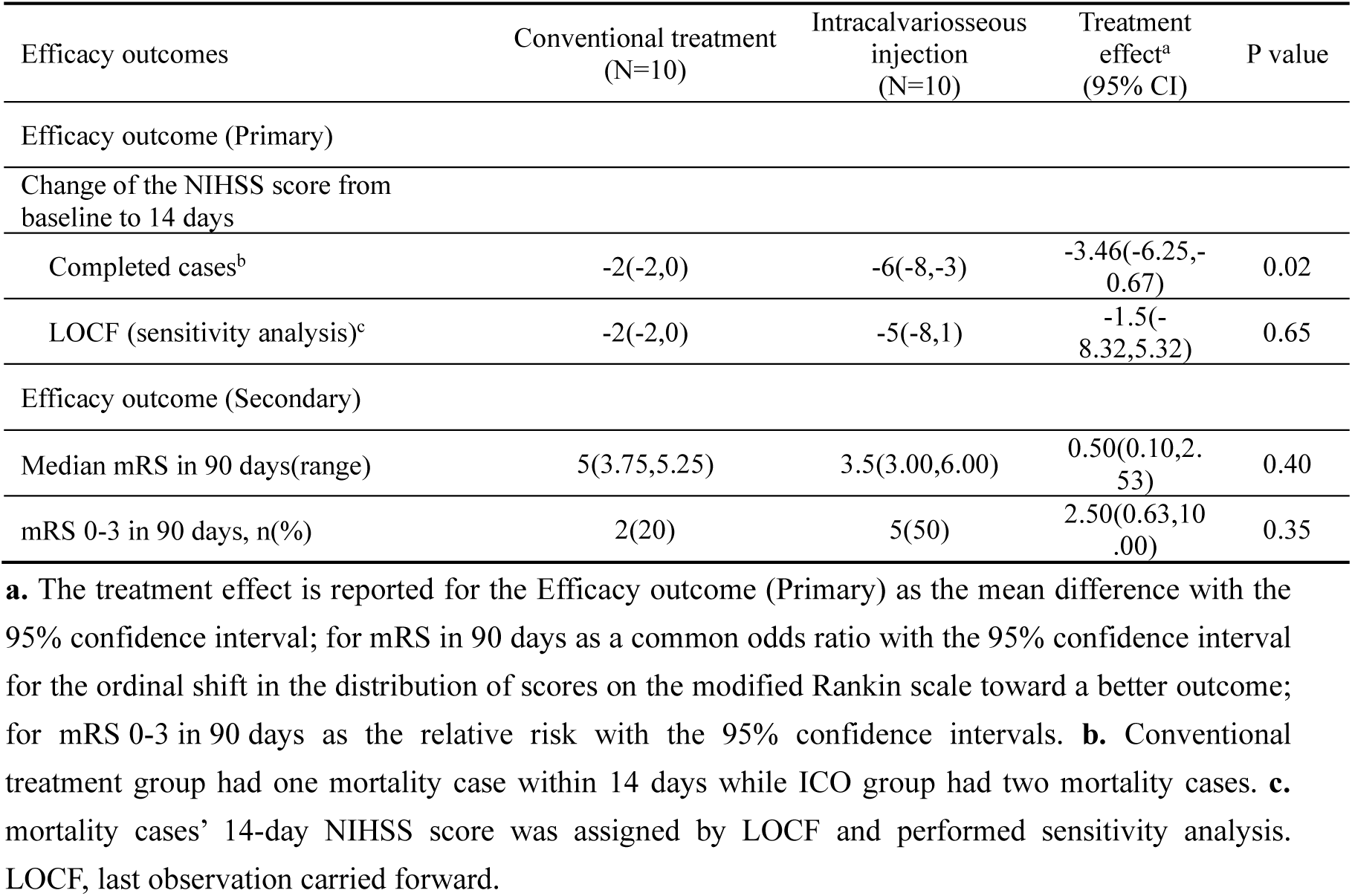
Primary efficacy outcomes.

| Efficacy outcomes | Conventional treatment<br>(N=10) | Intracalvariosseous<br>injection<br>(N=10) | Treatment<br>effect<sup>a</sup><br>(95% CI) | P value |
| --- | --- | --- | --- | --- |
| Efficacy outcome (Primary) |  |  |  |  |
| Change of the NIHSS score from<br>baseline to 14 days |  |  |  |  |
| Completed cases<sup>b</sup> | -2(-2,0) | -6(-8,-3) | -3.46(-6.25,-<br>0.67) | 0.02 |
| LOCF (sensitivity analysis)<sup>c</sup> | -2(-2,0) | -5(-8,1) | -1.5(-<br>8.32,5.32) | 0.65 |
| Efficacy outcome (Secondary) |  |  |  |  |
| Median mRS in 90 days(range) | 5(3.75,5.25) | 3.5(3.00,6.00) | 0.50(0.10,2.<br>53) | 0.40 |
| mRS 0-3 in 90 days, n(%) | 2(20) | 5(50) | 2.50(0.63,10<br>.00) | 0.35 |

**a.** The treatment effect is reported for the Efficacy outcome (Primary) as the mean difference with the 95% confidence interval; for mRS in 90 days as a common odds ratio with the 95% confidence interval for the ordinal shift in the distribution of scores on the modified Rankin scale toward a better outcome; for mRS 0-3 in 90 days as the relative risk with the 95% confidence intervals. **b.** Conventional treatment group had one mortality case within 14 days while ICO group had two mortality cases. **c.** mortality cases' 14-day NIHSS score was assigned by LOCF and performed sensitivity analysis. LOCF, last observation carried forward.

One case in the ICO group was diagnosed with meningitis at 75 days after symptoms onset and deemed to be secondary to decompressive craniectomy by the clinical event committee. Symptomatic and non-symptomatic intracalvarial hemorrhage was reported in 7 cases of the conventional treatment group and 4 in the ICO group (relative risk, 0.57 95% CI, 0.24 to 1.35; P=0.37) (Table 1), which might relate to the higher number of cardiogenic embolism and thrombolysis cases in the conventional treatment group. Within 90 days after disease onset, two fatal cases were reported in the conventional treatment group: one each of fatal stroke and ventricular fibrillation. Three fatal cases were reported in the ICO group with one each of fatal stroke, posterior circulation infarction and acute myocardial infarction. No significant differences between the two groups were observed in hepatic insufficiency, renal insufficiency, and mortality within 90 days. No cases of moderate to severe bleeding (defined by the GUSTO) or severe anemia were reported (Table 1).

### Secondary outcomes: efficacy of ICO in stroke patients

In secondary analyses up to day 14, the ICO group exhibited a greater reduction in NIHSS score compared to the conventional treatment group (median −6vs-2, P=0.02). Since NIHSS score is not applicable to deceased patients, a Last Observation Carries Forward (LOCF) sensitivity analysis was performed, which showed no statistically significant reduction in 14-day NIHSS score in the ICO group compared to baseline (median −5vs-2, P=0.65) (Table 2 and Fig.5 a-b). A tendency of shift in the distribution of scores on the modified Rankin scale at 90 days toward better outcomes was observed in favor of ICO injection over conventional treatment (Table 2 and Fig.5 c). Ten percent of patients experienced a reduction in NIHSS score greater than 4 points at the 14-day mark in the conventional treatment group, while in the ICO group, this proportion was notably higher at 60% (relative risk, 6; 95% CI,0.87 to 41.21; P=0.057). The rate of DC within 90 days was 40% in the conventional treatment group and 20% in the ICO group (relative risk, 0.5; 95% CI, 0.12 to 2.14; P=0.63) (Extended Table 3).

**Fig.5.**
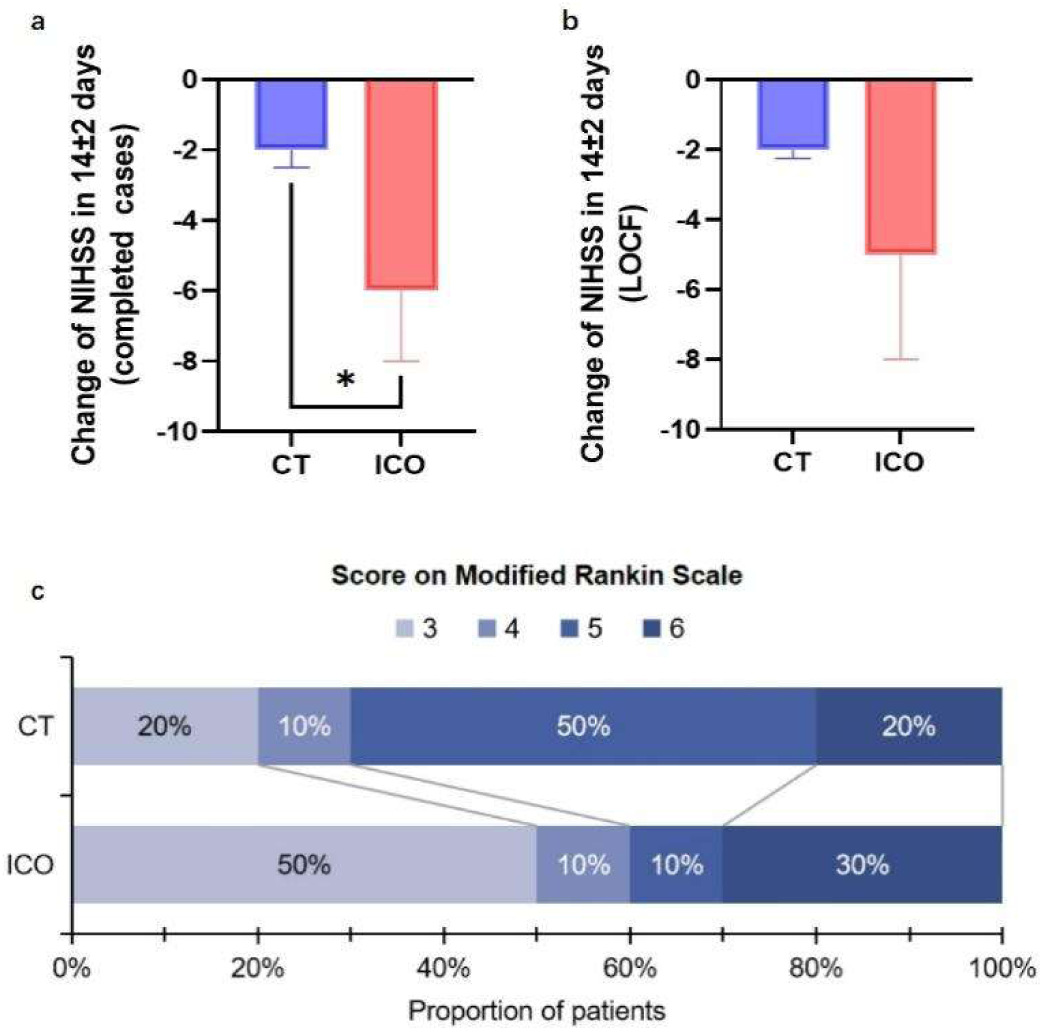
Bar Graph of efficacy outcomes. **a.** Median and interquartile range of change of NIHSS score in 14±2 days. **b.** Median and interquartile range of change of NIHSS score in 14±2 days, mortality cases’ 14-day NIHSS score was assigned by LOCF. **c.** Score on the modified Rankin Scale (mRS) at 90±7 days after onset of the symptoms among patients. NIHSS score was calculated using the Mann-Whitney U test * P< 0.05. CT, Conventional treatment; ICO, Intracalvariosseous injection; LOCF, last observation carried forward.

## Discussions

The discovery of microchannels and substance exchange between the skull and the meninges has made it possible to delivery drugs into the brain by intracalvariosseous injection. We optimized ICO injection with a single-hole injection which reduced skull damage and so was beneficial for maintaining the CNS immune environment as the skull is considered a myeloid-derived cell bank of brain parenchyma^25,26^. Considering that ICO injection requires local removal of a small portion of the skull, which is an invasive operation and may cause cranial bone marrow infection, we confirmed that ICO injection was safe and did not cause osteomyelitis determined by neutrophil cell number in the skull bone marrow and peripheral blood. Further, ICO injection in mice did not affect BBB permeability for molecules in the weight range of 4k Da-150k Da assessed using FITC-dextran. To explore feasibility and safety of ICO in clinical applications, the clinical trial SOLUTION was performed and demonstrated that outer skull plate drilling and ICO could be performed safely at the bedside in the ICU using standard neurosurgical instruments. The operation could be completed in 40 minutes and within 5 hours after hospital arrival. Compared to the conventional treatment group, the ICO group did not experience any severe adverse events related to the operative procedure, including skull penetration and intracalvarial bone marrow infection, supporting initial safety. In contrast to the rodents, the intracalvarial space of humans was much larger and enabled injection of enough drug solution without observable drug leakage. These outcomes suggested the potential for future clinical applications of the technology.

The lower rate for successful CNS drug development is due to, at least in part, the CNS accessibility. To elevate the exposure of drugs to brain tissue, it is necessary to increase the overall drug dose through intravenous injection, which undoubtedly increases drug toxicity in the periphery. In this study, the drug dosage of ICO injection of NA-1 or Y-3 is only 50% or 2.5% respectively as compared with IV injection, however, the observation of intraneuronal cytoplasmic FITC-NA1/Y-3 in brain and HPLC-detected Y-3 in brain confirmed that ICO injection can deliver the drug to the brain parenchyma even neurons. Therefore, ICO injection achieves high intracranial exposure with a lower drug dose. For drugs that cause peripheral toxicity due to IV injection with a high-dose, ICO injection may be a new injection route to reducing dosage, alleviating peripheral toxicity, and decreasing medical costs. Additionally, ICO injection enhances drug accessibility in the CNS by bypassing the BBB, which is a departure from previous studies that focused on BBB opening to enhance drug penetration. ICO injection does not impact the permeability of the BBB; instead, it facilitates drug delivery to the perivascular space via the microchannel between the skull and the dura mater, enabling drug entry into the brain.

Our study has several limitations. Firstly, despite apparent safety in both a stroke model and a small human cohort, ICO is an invasive approach. Second, CSF movement is influenced by factors such as sleep patterns, arterial pressure fluctuations, and the presence of different CNS pathologies^27,28^, which may impact the function of meningeal lymphatic vessels. Consequently, further validation of the effectiveness of ICO administration across diverse physiological and pathological conditions is warranted. Third, our findings indicated that following ICO injection, drug distribution primarily filled the skull marrow cavity and then enter brain, but a significant proportion of the drug retained within the skull. The efficacy of NA-1 did not increase with higher doses, possibly because the elevated concentration of NA-1 leads to a significant proportion of NA-1 remaining in the skull, thereby reducing drug entry efficiency into the brain parenchyma. Therefore, future research endeavors should focus on strategies to minimize drug residue and mitigate its potential impact on the skull bone marrow. Fourth, our application of ICO to inject Y-3 in patients with stroke suggested it might alleviate neurological impairment in patients within 14 days but could not reduce patient mortality. This might be related to the fact that patients in the conventional treatment group had larger hypoperfusion areas and prolonged interval between disease onset and enrollment into the treatment or may be a chance finding given the small sample size of the trial. Demonstration of efficacy requires confirmation in a large trial, and the current results should be considered as the combined effect of the injection method and the drug. Last, it is important to acknowledge that stroke itself may compromise BBB integrity, this potentially allowing a limited amount of drug to enter the brain parenchyma 24 hours post-stroke, though ICO itself could not influence BBB integrity.

In summary, the improved ICO injection is a novel drug delivery method that bypasses the BBB to achieve drug CNS accessibility in a short time. This study investigated the safety, feasibility and efficacy of ICO injection under both normal physiological conditions and in the context of ischemic stroke, which provided a foundation for the clinical translation of ICO injection. Moving forward, ICO injection could broaden candidates for new drug formulations in neurological condition and so provide a new choice for CNS drug therapy, and our subsequent research will aim to expand the scope by investigating the administration of various drug types via ICO and assessing their feasibility, safety, and efficacy across a spectrum of CNS diseases, including acute conditions such as traumatic brain injury and infection, as well as chronic diseases such as tumors and Alzheimer’s disease.

## Data Availability

All data produced in the present work are contained in the manuscript.

## Methods

### Animals

C57BL/6J mice and SD rats were purchased from Vitalriver Co., Ltd. Male mice at age of 8-10 weeks and male rats at age of 7-8 weeks were used in this study. Animals were housed in a controlled environment with consistent temperature (23±1°C) and humidity (50-60%). They were provided with unlimited access to food and water. All procedures in this study were in accordance with the Animal Care and Use Committees guidelines set by Capital Medical University.

### ICO injection

Mice and rats were anesthetized using nasal inhalation of isoflurane at a concentration of 1-1.5% and a flow rate of 0.5 ml/min. Animals were then securely positioned in a stereotaxic instrument, and their heads were carefully shaved and disinfected with iodophor. Subsequently, the occipital bone was exposed. Using a skull drill, holes were created in the occipital bone, ensuring penetration of the outer bone plate, while the inner bone cortex remained intact. The diameter of the hole was 1 mm for mice and 3 mm for rats. A micro syringe was connected to a flat-headed injection needle (Reward, 62203). Needle size was 30 G for mice and 26 G for rats. Needles were inserted into the exposed bone marrow barrier through the side hole, and microinjection (Gaoge, G019206) was initiated.

### Evans blue injection and concentration or fluorescence intensity detection

After injecting 5 μl of 10% Evans blue (EB) (Sigma, E2129) solution into the mice either by ICO or IV, 500 μl of 2.5% tribromoethanol-tert-amyl alcohol (Nanjing Aibei Biotechnology, M2920) solution was intraperitoneally administered for anesthesia at 10 min, 30 min, and 60 min. Eye extraction was used for blood collection and separation of plasma. This procedure was followed by cardiac perfusion with ice-cold PBS to remove blood. Skull, dura mater, and brain were further harvested separately with the brain microscopically dissected into cortex, striatum, and hippocampus. EB was extracted/dissolved from the minced tissues using 50% trichloroacetic acid (Sigma, T0699), and the absorbance of solution was measured at 620 nm using a microplate reader. Different concentrations of EB solution were used as standard solutions to calculate the concentration of EB in the test samples. Additionally, after the mice were injected with 5 μl of 10% EB solution through the skull bone marrow, they were anesthetized at 0.5 h, 1 h, 2 h, 6 h, 12 h, 24 h, and 72 h by intraperitoneal injections of 500 μl of 2.5% tribromoethanol-tert-amyl alcohol solution. The brain tissues were then immersed in a 4% paraformaldehyde (PFA) solution (Aladin, C104188), dehydrated using 30% sucrose (BioFroxx, 1245GR500), and subsequently frozen and sectioned (with a section thickness of 20 μm). Whole-brain scan was conducted using Texas red pathway.

### pMCAO animal model

8-10-week aged mice (21-25g) or rats (280-330 g) were anesthetized using inhalation of isoflurane at a concentration of 1-1.5% and a flow rate of 0.5 ml/min. With animal lying supine, the skin and muscle in the middle of the neck were cut to expose blood vessels. The right external carotid artery and right common carotid artery were ligated. A small cut was made superior to the ligation site of the right common carotid artery. A nylon suture bolt (Reward, Rat: MSRC40B200PK50, Mouse: MSMC21B120PK50) was inserted into the middle cerebral artery. The sham operation involved separating nerves and blood vessels but without inserting the suture. The skin was sutured layer by layer and the vital signs of the mice or rats were continuously monitored throughout the procedure^29^.

### Longa neurological function evaluation

One hour after inducing the pMCAO model, the Longa method was used to perform an initial evaluation of neurological function of the mice or rats to determine if the model was successful. A score of 2-3 indicated a successful pMCAO model^30^.

### FITC-dextran injection

Mice were injected with 5 μl of 10% EB solution by ICO 24 hours after sham or pMCAO model induction. After 1 hour, the mice were intravenously injected with 100 μl of 4k Da FITC-dextran (Sigma, 68059) (50 mg/ml), 40k Da FITC-dextran (Sigma, 74817) (20 mg/ml), and 150k Da FITC-dextran (Sigma, 53379) (20 mg/ml). After 10 minutes of blood circulation, mouse brain tissues from the 40k Da FITC-dextran and 150k Da FITC-dextran groups were immersed in 4% PFA for 12 hours and then sectioned into 50 μm frozen slices. The brain tissues from the 4k Da FITC-dextran group were directly embedded in OCT compound and sectioned into 50 μm frozen slices^31^.

### Flow cytometry

After 24 hours of pMCAO induction, the mice were sacrificed, and their blood was collected from the retroorbital sinus with an anticoagulant to prevent blood clotting. After PBS perfusion through the heart, the skull was removed from the body and placed in 1X PBS solution after washing-off surface blood and muscle, then filtering through a 100 μm filter membrane to obtain a suspension of skull marrow cells. A mixture of blood and skull marrow cell suspension were treated with 1X red blood cell lysis buffer (BD, 555899) to remove red blood cells, and cell precipitate was obtained after centrifugation. Then the cell was resuspended and incubated with FITC-CD45 antibody (eBioscience, 103108, 1:200), PB450-Ly6G antibody (BD, 746448, 1:200), and APC-CD11b antibody (eBioscience, 17011282, 1:200) for 1 hour to identify neutrophils in the skull marrow and peripheral blood. The quantity of neutrophils in the skull marrow and peripheral blood was measured using a flow cytometer (Beckman, cytoflex).

### Transparency processing of skull-dura-brain integrated tissue

After injecting 5 μl of Cy3-labeled NA1 (Cy3-NA1) into the skull bone marrow, 10 μl of 488-labeled rabbit polyclonal Lyve1 antibody (R&D, FAB2125G) was immediately injected through the cisterna magna at a speed of 2 μl/min. Additionally, 647-labeled mouse monoclonal CD31 antibody (Abcam, ab305210, diluted 1:5 in physiological saline) was injected through the tail vein. After allowing the antibodies to circulate for 1 hour, brain tissue was collected after PBS perfusion. The fixed tissue was then soaked in PBS solution following 12 hours of fixation with PFA. Tissue transparency was performed using the FDISCO in vitro optical transparency reagent (Javisbio Co., Ltd.) following the provided instructions.

### PSD95 inhibitor NA-1/Y-3 injection by ICO

To determine the optimal drug dosage for ICO injection of NA-1/Y-3 in the treatment of stroke, rats were randomly assigned to NA-1 or Y-3 drug groups. One hour after the pMCAO model, rats with a Longa score of 2-3 were administered drug injection. In the NA-1 group, rats were randomly assigned to the control group, IV group, or ICO A-C groups. The IV group received NA-1 (3 mM, 7.8 mg/kg) via tail vein injection, while ICO A, B, and C groups received NA-1 solution 5 mM (1.95 mg/kg), 10 Mm (3.9 mg/kg), and 20 mM (7.8 mg/kg) respectively, via ICO injection. NA-1 was synthesized by Peptide Biotech Co., Ltd (China. Beijing). The Y-3 group was randomly assigned to the control group, IV group, or ICO A-F groups. The IV group received Y-3 (2 mg/ml, 4 mg/kg) via tail vein injection, while ICO A-F groups received Y-3 at doses of 0.2 mg/kg, 0.1 mg/kg, 0.05 mg/kg, 0.025 mg/kg, 0.0125 mg/kg, and 0.0033 mg/kg respectively. Y-3 was provided by Neurodawn Pharmaceutical Co., Ltd (China. Nanjing).

Efficacy of the PSD95 inhibitor NA-1/Y-3 after ICO injection in the pMCAO model was than assessed at 24 hours and 7 days. Rats were randomly assigned to sham, control, NA1-IV, NA1-ICO, NA1-ICO-Vehicle, Y3-IV, Y3-ICO, and Y3-ICO-Vehicle groups. In addition to the sham and control group, the other groups received drug treatment 1 hour after pMCAO model. The NA1-IV group received NA-1 solution (3 mM, 7.8 mg/kg) via tail vein injection, while the NA1-ICO group received NA-1 solution (10 mM, 3.9 mg/kg) via ICO injection, and the ICO-Vehicle group received an AA solution (10 mM, 3.9 mg/kg) via ICO injection. AA has a different amino acid sequence with NA-1^32^, was synthesized by Peptide Biotech Ltd. The NA1-IV, NA1-ICO, and NA1-ICO-Vehicle group received a single dose. The Y3-IV group received Y-3 (2 mg/ml, 4 mg/kg) via tail vein injection, while the Y3-ICO group received Y-3 solution (2 mg/ml, 0.1 mg/kg) via ICO injection, and the Y3-ICO-Vehicle group received Y-3 solvent (0.1 ml/kg) via ICO injection, Y-3 solvent provided by Neurodawn Pharmaceutical Co., Ltd (China. Nanjing). The Y3-IV, Y3-ICO, and Y3-ICO-Vehicle group received continuous injections for 3 days. Rats were sacrificed at 24 hours and 7 days after pMCAO induction, and brain tissue was immersed in 4% PFA solution, dehydrated in 30% sucrose solution, and used for frozen sectioning.

To assess FITC-labeled NA-1/Y-3 tracking, rats were randomly assigned to NA1-IV, NA1-ICO, Y3-IV, and Y3-ICO groups. One hour after pMCAO induction, rats with Longa scores of 2-3 were injected with FITC-NA1 (3 mM, 7.8 mg/kg)/FITC-Y3 (2 mg/ml, 4 mg/kg) via tail vein in the NA1-IV/Y3-IV group, and with FITC-NA1 (10 mM, 3.9 mg/kg)/FITC-Y3 (2 mg/ml, 0.1 mg/kg) via ICO injection in the NA1-ICO/Y3-ICO group. After 24 hours, the rats were sacrificed, and brain tissue was fixed in 4% PFA, dehydrated in 30% sucrose, and frozen sectioned into 20 μm slices.

### Triphenyl tetrazolium chloride staining

First, the brain tissue was placed into a rat brain mold. Then, brain tissue was sliced into coronal slices with thickness of 2 mm. Next, slices were immersed in a 2% TTC (Sigma, T8879) solution and soaked in the solution at a temperature of 37°C for 10 minutes. Finally, an EPSON scanner was used to scan the stained brain slices.

### Infarct area calculating

The total infarct area is the sum of the infarct area of all brain slices, and the infarct area of each brain slice based on the formula: Infarct area=Contralateral brain area-ipsilateral infarct brain area.

### Neurological functional assessment

Rats were assessed for their neurological function by researchers who were given unidentified animals. The modified version of neurological deficit assessment (mNSS) was used to evaluate the neurological function of the rats, with a total score of 18 points^33^. A higher score indicated a more severe neurological impairment.

### Rotarod test

The Rotarod experiment was conducted by an experimenter blinded to experimental group. Rats in the pMCAO model underwent Rotarod preconditioning for three consecutive days, with the aim of enabling the rats to remain on the instrument for a minimum of five minutes. Subsequently, Rotarod testing was carried out on the 1st, 3rd, 5th, and 7th days following the model. Each rat was subjected to three tests on the same day, with a 15-minute interval between each test.

### Immunofluorescence staining

Before performing immunofluorescence staining, brain slices were allowed to return to room temperature and the OCT glue present on the slices was dissolved. To rupture the membranes, slices were incubated for 50 minutes in a 1% PBST solution (Triton-100X dissolved in PBS). Following these slices were blocked with a blocking solution (Beyotime, P0102) for 1 hour. For the staining, slices were incubated overnidght at 4°C with the following primary antibodies: Rabbit polyclonal NueN antibody (Millipore, ABN90, 1:500), Mouse monoclonal nNOS antibody (Santa Cruz Biotechnology, Sc-5302, 1:100), and Mouse monoclonal Caspase3 antibody (Proteintech, 6470-2-Ig, 1:200). After incubation, the slices were washed and incubated at room temperature for 1 hour with appropriate secondary antibodies: Goat anti-Rabbit 594 (Abcam, ab150080, 1:500), Goat anti-Mouse 488 (Jacksonlab, 715545151, 1:500), and Goat anti-Mouse Cy3 (Jacksonlab, 715165151, 1:500). Finally, the slices were washed again and mounted them using DAPI mounting medium (Solarbio, S2110).

### Detection of Y3 concentration in the tissue

At 1 hour, 2 hours, and 4 hours after injecting Y-3 into rats with pMCAO model, tissue samples were taken. The obtained sample tissues included plasma, skull, dura mater, CSF, cerebellum, cerebral cortex, striatum, hippocampus, and deep cervical lymph nodes (CLN). All tissues underwent cardiac perfusion to remove blood before acquisition. Y-3 was extracted from the skull using 50% methanol water homogenate, while 20% methanol water was used for the tissues. The concentration of Y-3 in each tissue homogenate was detected by high performance liquid chromatography (HPLC). The tissue to blood partition coefficients (*K*p) was used to represent the ratio of drug concentration in tissues to drug concentration in plasma based on the formula: *K*p=AUC tissue(0-t)/AUC plasma(0-t)^34^.

### Liver and kidney function tests

Liver function test indicators included AST (Aspartate Aminotransferase) and ALT (Alanine Aminotransferase). Kidney function test indicators included UREA and CREA (Creatinine). After anesthesia, blood samples were collected from the rat’s heart, placed in anticoagulant tubes, centrifuged at 4000 rpm for 15 minutes, and the supernatant was collected. The obtained rat plasma was then processed according to the requirements of AST, ALT, UREA, and CREA test kits (Mindray), and the values of each indicator were measured using a fully automatic biochemical analyzer (Mindray, BS-240Vet).

### Microscopy imaging

The whole brain slices were scanned using a Vectra Polaris fully automated imaging system (Perkin-Elmer) under a 20-time amplification microscope, and the exposure time was set according to different slices. We used a confocal microscope (ZEISS, 710) to scan at 20-time and 40-time amplification microscope, adjusting exposure time to obtain the best images.

### Imaging Data processing

Image J software (version 1.8.0) was used for analysis of infarct area, BBB leakage analysis^35^, fluorescence intensity analysis, and cell counting. 3D imaging analysis was performed using Imaris software (version 9.0.1). CytExpert software (version 2.1.0.92) was used for analysis of flow cytometry results.

### Study design and trial patients of clinical trial

SOLUTION was a prospective, randomized, open-labeled, blinded endpoint clinical trial. The study was conducted in Beijing Tiantan Hospital, China. The protocol was approved by the Institutional Review Board (IRB) of Beijing Tiantan Hospital (IRB Approval Number:KY2023-052-02), and the study was carried out in accordance with the principles of the Helsinki Declaration and the International Council for Harmonization of Good Clinical Practice guidelines. A steering committee was responsible for the design and implementation of the trial, while a data and safety monitoring committee oversaw the trial and conducted regular safety assessments. The data was analyzed by the data management center of the Chinese Neurological Clinical Research Center. All patients or their legal representatives were informed about the study protocol and provided written informed consent prior to enrollment. The trial staff ensured the accuracy, completeness of the data, and the fidelity of the trial to the protocol. This trial was registered on ClinicalTrials.gov, NCT05849805.

The trial enrolled patients aged 18-75 years without gender restriction. Operations had to be completed within 24 hours. The National Institutes of Health Stroke Scale (NIHSS, ranged from 0 to 42, higher scores indicating more severe neurological deficits) score of 16 to 30 was required for entry.

The core infarction volume (defined as relative cerebral blood flow [rCBF] <30% volume in CT perfusion [CTP]) encompassed more than a half of the MCA territory or an Alberta Stroke Program Early CT Score (ASPECTS, a scale from 0-10 to measure infarct size, with lower scores reflecting larger infarcts) of ≤6. Participants undergoing reperfusion therapy with poor treatment outcomes were eligible (defined as deterioration or the improvement of post-treatment NIHSS ≤4 or deteriorated, and the total NIHSS score remained between 16 to 30). Additionally, patients all shared a pre-stroke modified Rankin Scale (mRS) score ≤1. (The scale ranging from 0-6 assesses capacity for daily activities, with higher scores indicating greater disability. A score of 6 signifies death.)

The primary exclusion criteria included: presence of acute cerebral hemorrhage or subarachnoid hemorrhage; acute posterior circulation ischemia; bilateral pupil fixation or pupillary reflex disappeared; DC was scheduled before randomization; contraindications for intracalvarial drug administration, e.g. skull fracture, skull infection, subdural/external hematoma, sub-scalp hematoma, scalp, or subcutaneous infection. A full list of inclusion and exclusion criteria is available in the supplemental Table 2.

### Randomization and Interventions of clinical trial

Patients with potential mMCAI were continuously screened in the emergency room (ER). Those meeting the inclusion criteria were randomly assigned to the Intracalvariosseous Injection (ICO) group or the Conventional Treatment (CT) group at a 1:1 ratio through simple random allocation. The randomization code was obtained by the random-permuted fixed-size blocks methods from the Statistics and Data Centre at the China National Clinical Research Centre for Neurological Diseases. All the patients received active treatment and care according to the Chinese Clinical Management Guidelines for Cerebrovascular Diseases (2nd Edition). In addition, the ICO group received an ICO Y-3 injection, with a dosage calculated as 32 μg/kg. Referring to stereotactic aspiration^36^, we designed a specific operational protocol (details of protocol and operational instruments are in Supplementary 3). All the participants were admitted to the intensive care unit (ICU) to monitor for the occurrence of malignant cerebral edema. Operations in the ICO group were carried out under the monitoring, with salvage equipment and specific medical personnel on standby. During hospitalization, patients were regularly undergoing head CT scan (including bone windows), chest CT scan, and laboratory tests to monitor for potential infectious events and other adverse events. After patients were out of life-threatening conditions, they were transferred to general wards for rehabilitation training.

### Clinical trial outcomes

The primary outcomes of our study were set to evaluate feasibility and safety of this innovative protocol. Feasibility outcomes were defined as successful drilling through the skull internal plate, drug leakage during the injection, the patient refusal to continue, and failure for other reasons during our 3-day treatments. Safety outcomes included infection events (skin infection, osteomyelitis, or intracalvarial infection), symptomatic and asymptomatic intracalvarial hemorrhage, moderate to severe bleeding (defined by the GUSTO), hepatic insufficiency, renal insufficiency during the treatment, severe or extremely severe anemia (hemoglobin <60g / L), mortality, incidence of other adverse events/serious adverse events.

Secondary efficacy outcomes included: NIHSS score increase of ≥4 at 7±2 days compared to baseline; NIHSS motor score increase of ≥2 at 7±2 days compared to baseline; changes of core infarction volume from baseline to 7±2 days; NIHSS score increase of ≥4 at 14±2 days compared to baseline; NIHSS motor score increase of ≥2 at 14±2 days compared to baseline; changes in GCS scores from baseline values to 14±2 days or at discharge; rate of DC; mRS score 0-3 points at 90±7 days; days of NICU hospitalization; cost of the NICU hospitalization.

NIHSS score and mRS score were evaluated by two specialized neurologists. In instances of discordant assessment outcomes, adjudication was performed by a senior clinician to ensure the accuracy and reliability of the diagnostic evaluation. Prior to scoring, the physicians ensured that the heads of the patients in both groups were covered and appearance of dressings were consistent. Adverse events were adjudicated by a specialized clinical-event adjudication committee (CEC). The neurologists performing the scoring and the CEC members were blinded to the patients’ group assignments.

### Statistical Analysis

Animal experiments: GraphPad Prism 8.0.2 software was utilized for data analysis. Non-parametric testing was employed to determine if the data followed a normal distribution. Data that adhered to a normal distribution were presented as mean ± standard error of mean (SEM). The one-way NOVNA test was used for multiple group comparisons, and the two-way NOVNA test was used for analyses involving multiple factors and components, among them Tukey’s multiple comparisons test was used. Additionally, Survival analysis was conducted using Log-rank (Mantel Cox) test. A confidence interval of 95% was applied, and a p-value of less than 0.05 was considered statistically significant.

Clinical trial: As an initial exploratory study, we enrolled 20 patients. All numeric variables were analyzed by the Shapiro-Wilk test to determine whether they followed normal distributions. Normally distributed data are described as means ± standard deviations (SD), while non-normally distributed data were described as median (interquartile range, IQR) values. Categorical variables were expressed as counts and frequencies. Feasibility measures were only statistically described. All safety measures were compared between groups using Fisher’s exact test. The primary effectiveness measures were compared using the Mann-Whitney U test. Since NIHSS score and GCS score are not applicable for mortality cases, relevant score was treated as missing values and assigned by last observation carried forward (LOCF) which meant the values were assigned by the most recent valid scores. Among the secondary effectiveness measures, rates of NIHSS improvement, NIHSS motor limbs score improvement and mRS 0-3 in 90 days were compared using Fisher’s exact test. Differences in NICU hospitalization days and costs were compared using the Mann-Whitney U test. Statistical analyses were performed using SAS software, version 9.4 (SAS Institute). All statistical tests will be two-sided, with P <0.05 considered statistically significant.

## Data availability

Source data are provided with this paper.

## Acknowledgements

We thank Xuan Wang for analysis of clinical results; Miao Wen for patient’s management; Dongya Zhu, S Clay Johnston, Philip M Bath, and Ying Fu for feedback and discussions; Shibao Yang and the staff of Neurodawn Pharmaceutical Co., Ltd including Yao Hua and Sainan Lu for performing HPLC; and all of the members of the Yilong Wang laboratory for their contribution to the study. This work was supported by grants from The National Natural Science Foundation of China (No. 81825007); Beijing Outstanding Young Scientist Program (No. BJJWZYJH01201910025030); Capital special Funds for Health Improvement and Research (2022-2-2045); National Key R&D Program of China (2022YFF1501500, 2022YFF1501501, 2022YFF1501502, 2022YFF1501503, 2022YFF1501504, 2022YFF1501505); Youth Beijing Scholar Program (No.010); Beijing Laboratory of Oral Health (PXM2021_014226_000041); Beijing Talent Project: Innovation and Development (No. 2018A12); National Ten-Thousand Talent Plan”-Leadership of Scientific and Technological Innovation; National Key R&D Program of China (No. 2017YFC1307900, 2017YFC1307905).

## Author contributions

W. L. designed and performed the preclinical experiment, analyzed and interpreted data of animal experiment, created the figures and wrote the manuscript. N. W. designed and executed clinical trial, had full access to all the data in clinical trials, conducted the surgery, analyzed and interpreted data of clinical trial, created the figures and wrote the manuscript. M. Y. designed and executed clinical trial, had full access to all the data in clinical trials, assessed scale of clinal trial, interpreted data, revised and reviewed the manuscript. M. Z. assessed scale of clinal trial. X. L. assisted in the design of animal experiment. K. S. assisted in the design of animal experiment. W. L. conducted the surgery. W. O. conducted the surgery. L. L. treated and managed patients. Z. Y. treated and managed patients. Y. L. treated and managed patients. M. Z. designed clinical trial and assessed scale of clinal trial. X. L. treated and managed patients. H. Q. treated and managed patients. F.-D. S. provided intellectual contributions. Y. P. analyzed data. C. W. performed the preclinical experiment. X. F. assisted in the execution of clinical trial. X. Z. provided intellectual contributions in clinical trial. M. Z. assisted in the revision of manuscript. Y. W. provided resources and intellectual contribution. Y. W. Conceptualized and managed whole project, had full access to all the data in clinical trial, analyzed and interpreted data, made final approval of the article.

## Additional information

Supplementary Information is available for this paper.

Correspondence and requests for materials should be addressed to Wenqian Liu or Nanxing Wang or Yilong Wang.

---

**Extended Data Fig. 1.**
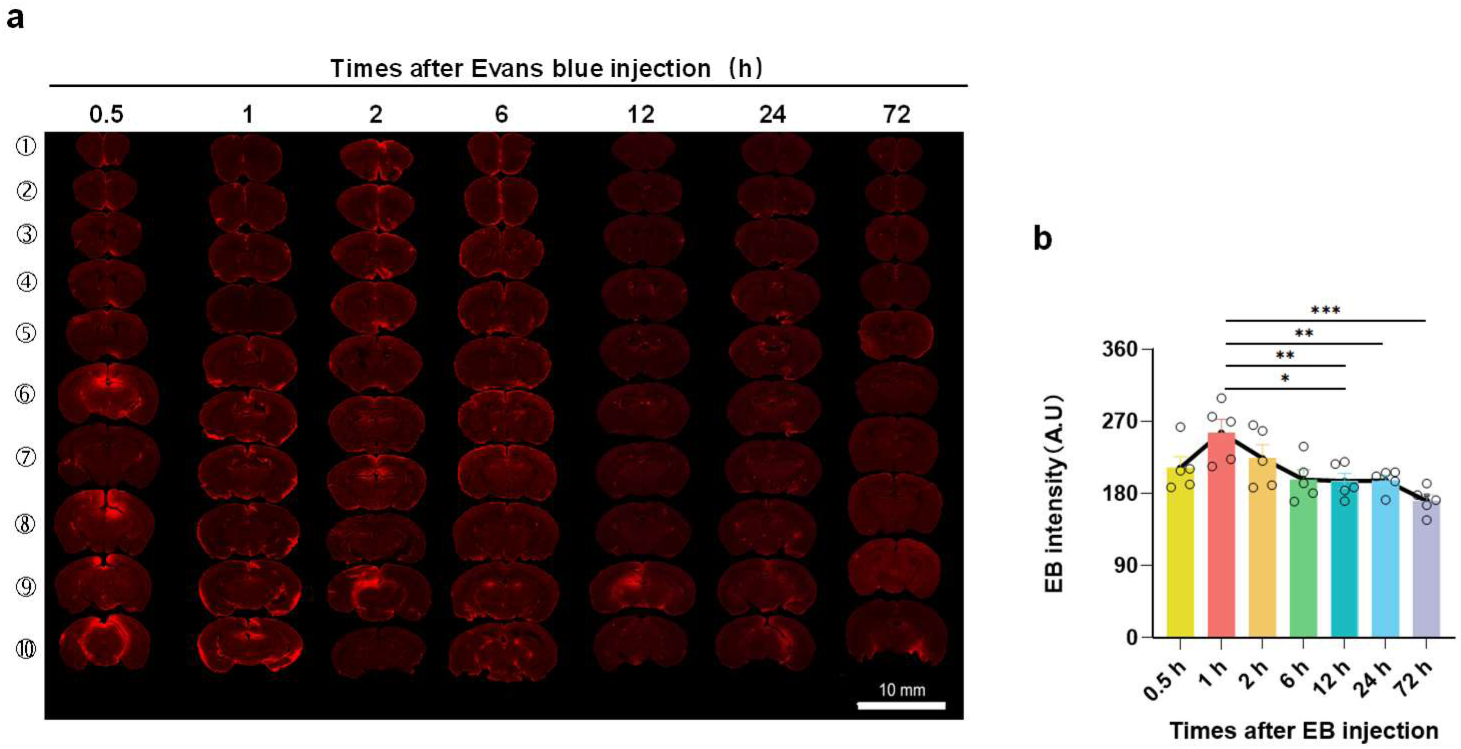
Metabolism and distribution of EB in brain tissues after ICO injections. **a-c.** Distribution of EB in brain tissues at 0.5 h, 1 h, 2 h, 6 h, 12 h, 24 h, and 72 h after the ICO injections of EB(*n*=5). (a) EB fluorescence intensity at different brain regions, scale=10 mm. (b)Statistical chart of EB fluorescence intensity in whole brain. Data are mean ± SEM. b. One-way ANOVA (Tukey’s multiple comparisons test). *p<0.05, **p<0.01, ***p<0.001.

**Extended Data Fig. 2.**
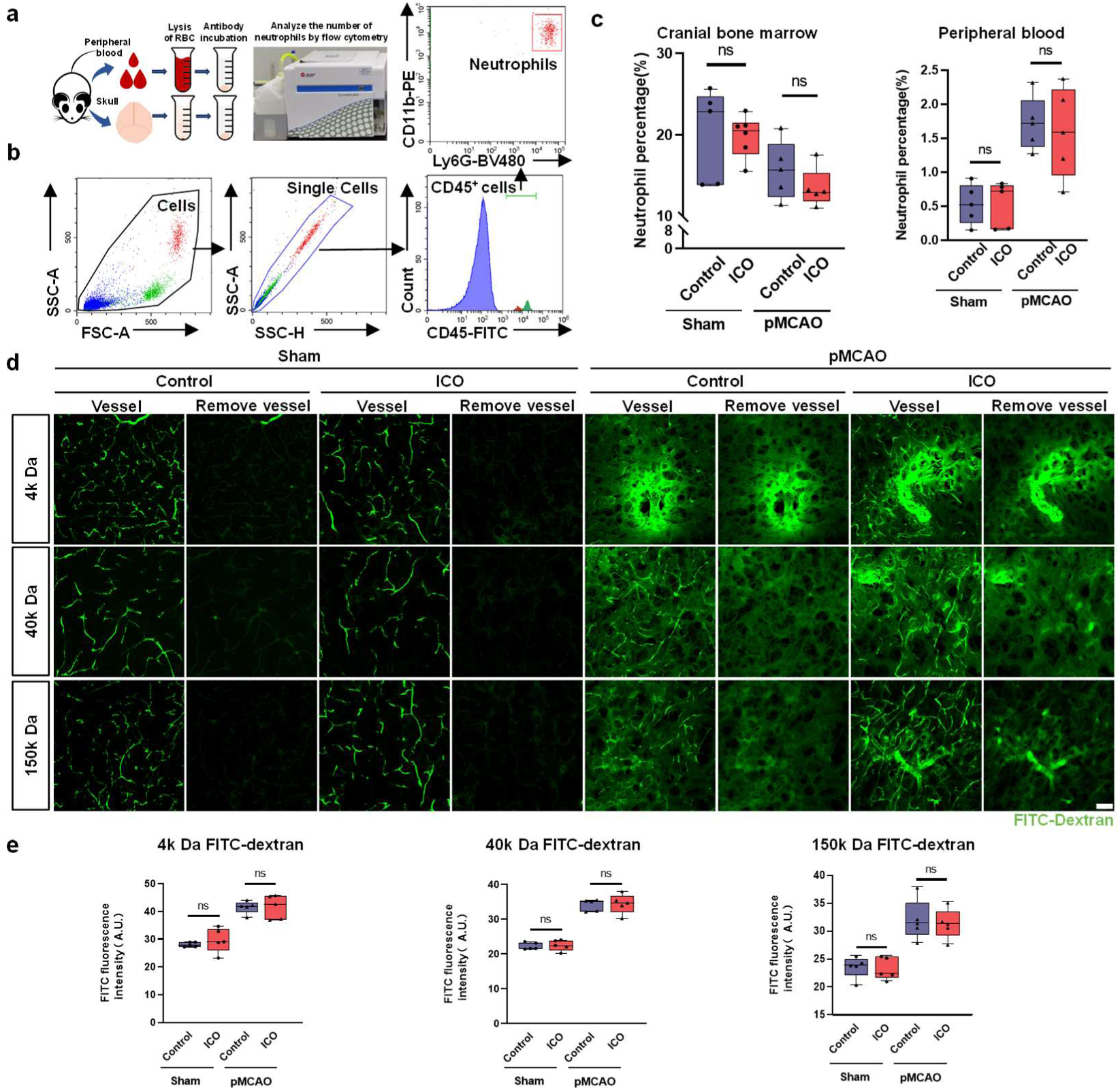
ICO injection does not cause skull bone marrow infection and changes in BBB permeability. **a.** Flow cytometry processing of peripheral blood and skull bone marrow cells. **b.** Flow cytometry gating strategy of neutrophil (CD45^+^CD11b^+^Ly6G^+^). c. Statistical chart of neutrophil ratio between cranial bone marrow (left panel) and peripheral blood (right panel) in each group(*n=5-6*). d. Capillary diagrams of different groups before and after removing the vessels by Image J, scale=50 μm. e. Leakage of FITC-Dextran with molecular weights of 4k Da, 40k Da, and 150k Da in vessels 1 hour after the ICO injection (*n=5-6*). Data are mean ± SEM. One-way ANOVA (Tukey’s multiple comparisons test). ns, no significance.

**Extended Data Fig. 3.**
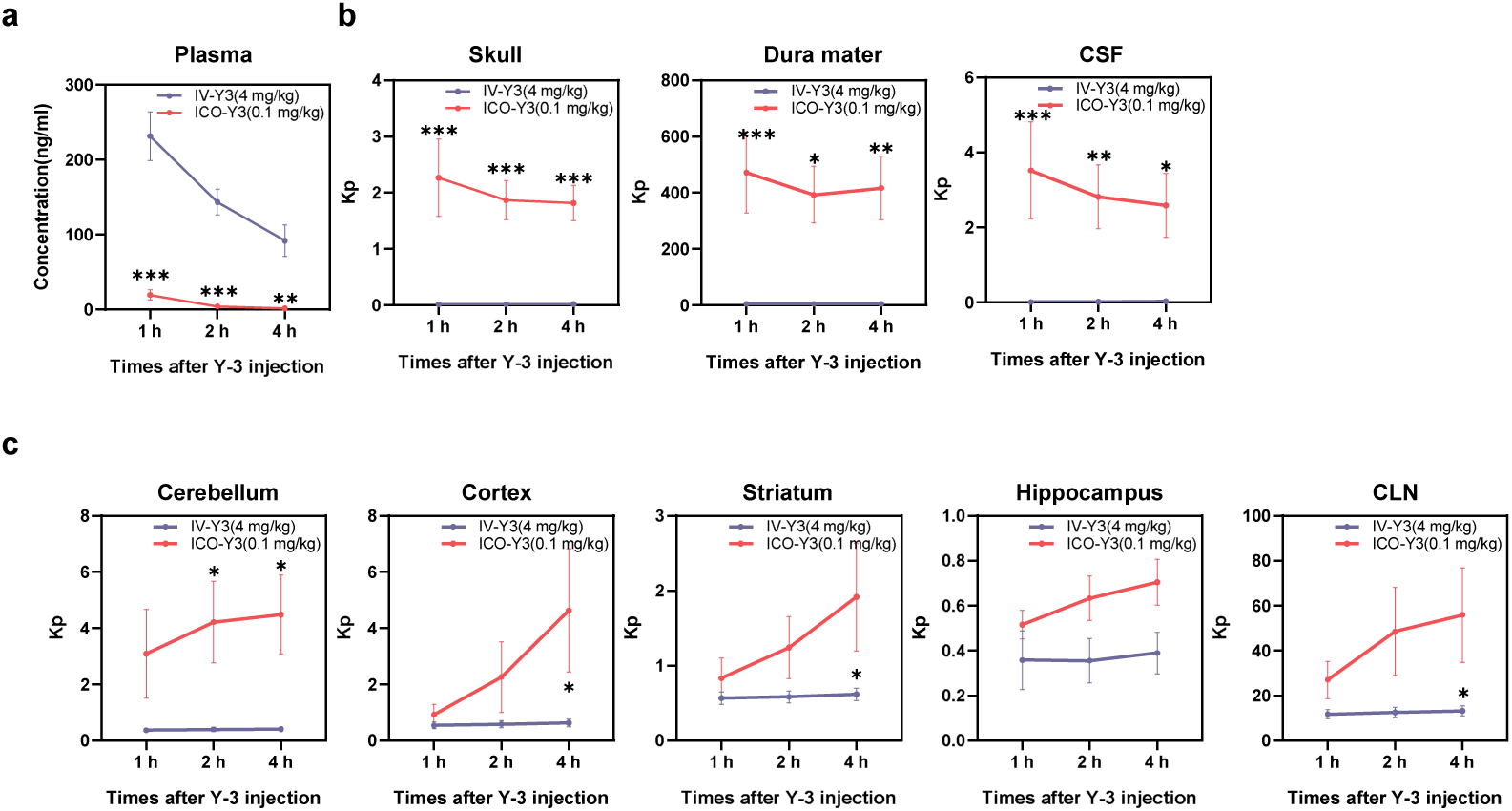
The concentration of Y-3 injected by IV or ICO after pMCAO model 1-4 hours in plasma and the *K*p of tissue. **a.** The concentration of Y-3 injected by IV or ICO in plasma(*n=5-6*). **b.** The *K*p with a deceased trend in skull, dura mater and CSF. c. The *K*p with an increased trend in cerebellum, cortex, striatum, hippocampus, and deep cervical lymph nodes (CLN). Data are mean ± SEM. Two-way ANOVA (Tukey’s multiple comparisons test). *p<0.05, **p<0.01, ***p<0.001.

**Extended Data Fig. 4.**
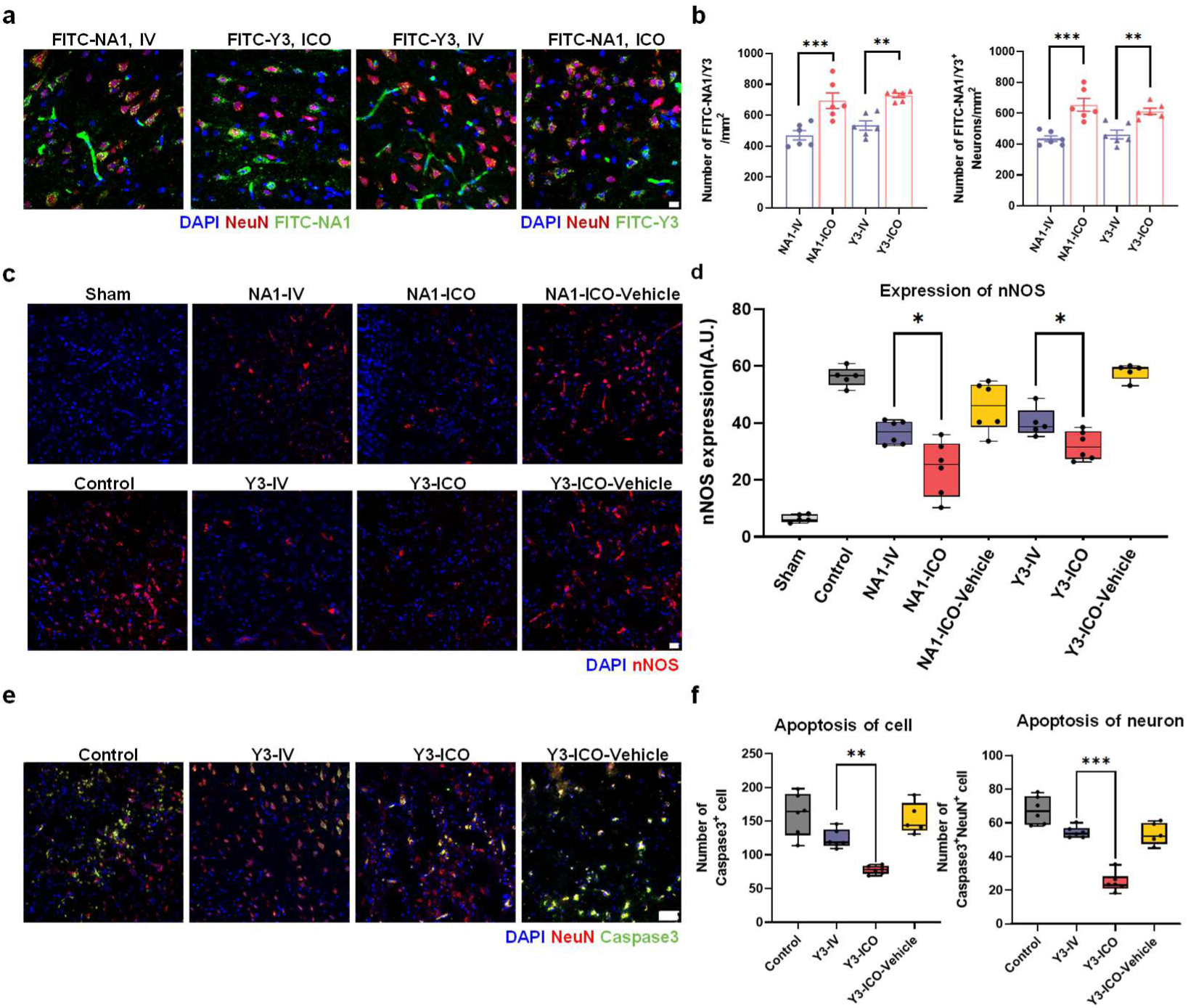
Quantification of NA-1/Y-3 drug levels in brain tissue and assessment of nNOS expression and cellular apoptosis following ICO injection in the pMCAO model. **a-b.** Neuronal uptake of FITC molecules 24 hours after IV or ICO injection of FITC-labeled NA-1/Y-3 (*n=5-6*). (a) Fluorescence images showed co-staining of FITC-NA1/Y3 with neurons in each group, scale=25 μm. (b) Quantification of FITC fluorescence molecules (left panel) and neurons with uptake of FITC molecules (right panel). **c-d.** Semi-quantitative analysis of nNOS fluorescence 24 hours after IV or ICO injection of NA-1/Y-3 (*n=5-6*). (c) Fluorescence staining images of nNOS in each group, scale=50 μm. (d) Statistical analysis of nNOS fluorescence intensity. e-f. Assessment of cellular apoptosis 7 days after ICO injection of Y-3 (*n=5-6*). (e) Co-staining of Caspase3 and NeuN fluorescence in each group, scale=50 μm. (f) Quantification of Caspase3-positive cells (left panel) and Caspase3 and NeuN double-positive cells (right panel). Data are mean ± SEM., b. One-way ANOVA (Tukey’s multiple comparisons test). Left Y axis, **p<0.01, ***p<0.001; Right Y axis, ##p<0.01, ###p<0.001. c. One-way ANOVA (Tukey’s multiple comparisons test). *p<0.05, **p<0.01, ***p<0.001.

**Extended Data Fig. 5.**
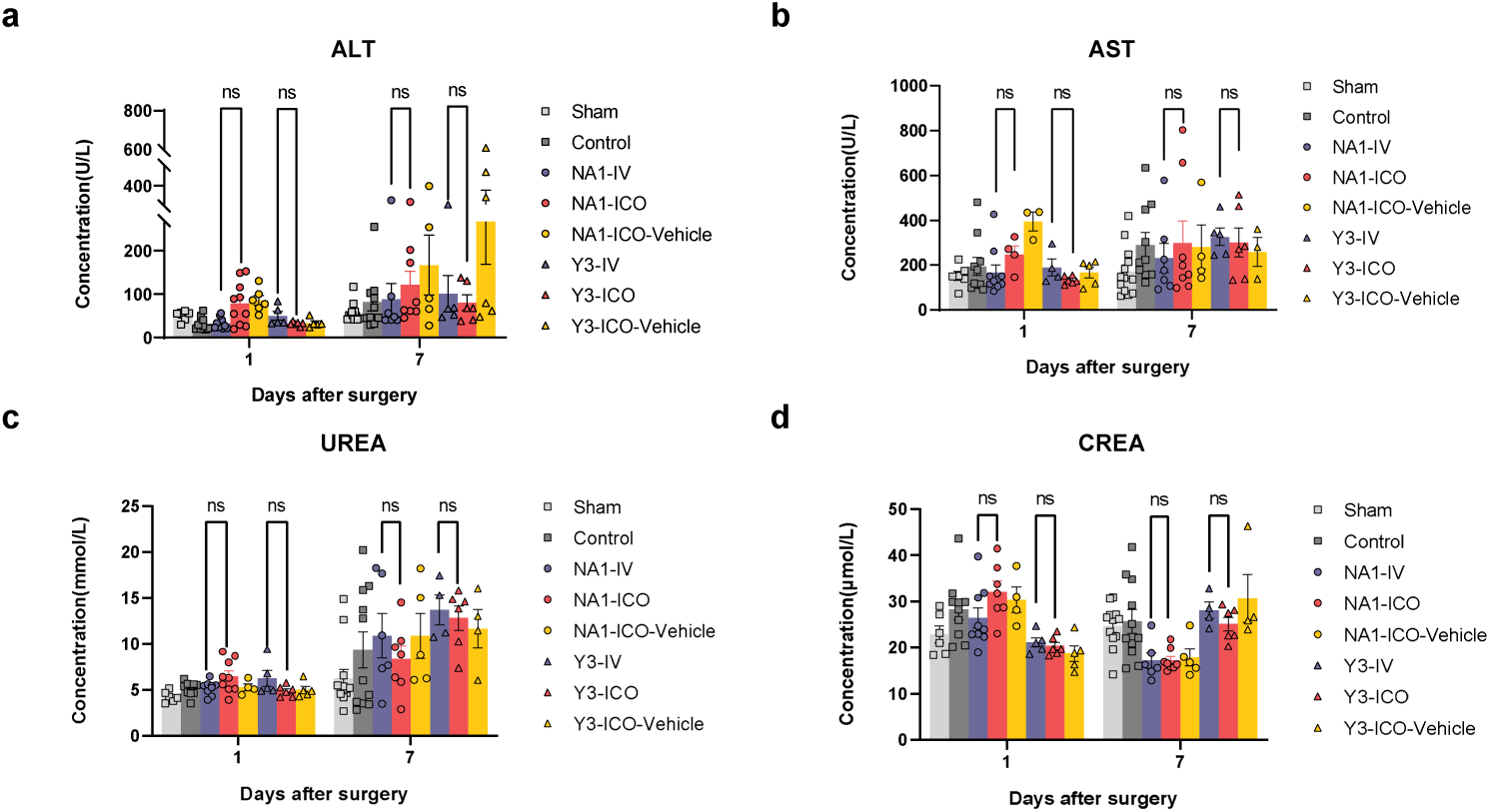
Differences in liver and kidney function after injections of NA-1/Y-3 by ICO or IV following pMCAO model. **a-b.** Statistical chart of liver function damage in each group, ALT (a) and AST (b), (*n=5-10*). **c-d.** Statistical chart of kidney function damage in each group, UREA (c) and CREA (d), (*n=5-10*). Data are mean ± SEM. Two-way ANOVA (Tukey’s multiple comparisons test). ns, no significance.

**Extended Data Fig. 6.**
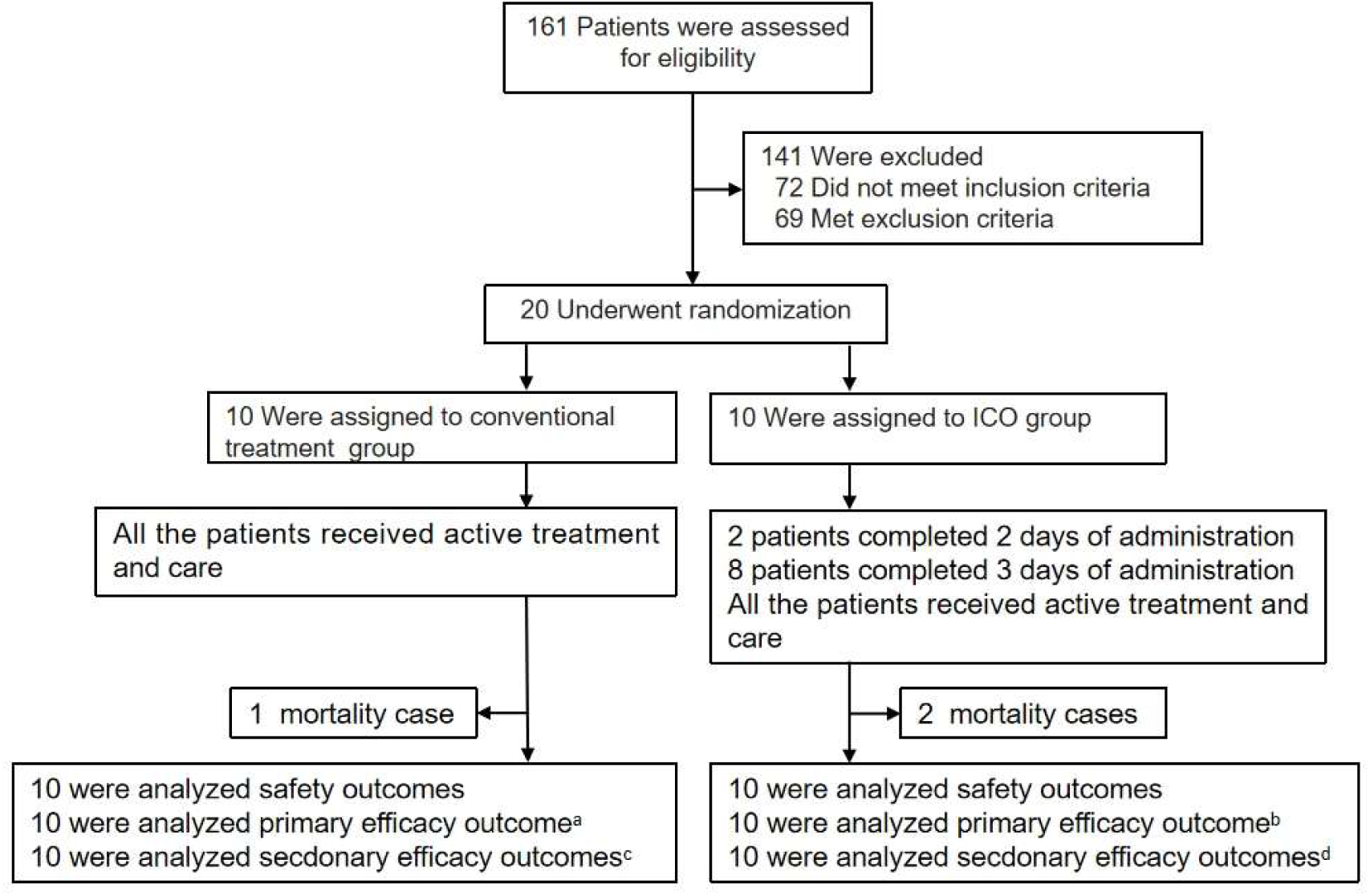
Enrollment and Randomization of Patients. **a-b.** Conventional treatment group had one mortality case and ICO group had two mortality cases. All the mortality case were treated as missing values and assigned by LOCF and performed sensitivity analysis. **c-d.** One patient in the Conventional treatment group requested early discharge from the hospital. Thus, each group had two patients that did not complete the 7-day CTP. CT, Conventional treatment; ICO, Intracalvariosseous injection; LOCF, last observation carried forward; CTP, computed tomography perfusion.

**Extended Data Table 1.**
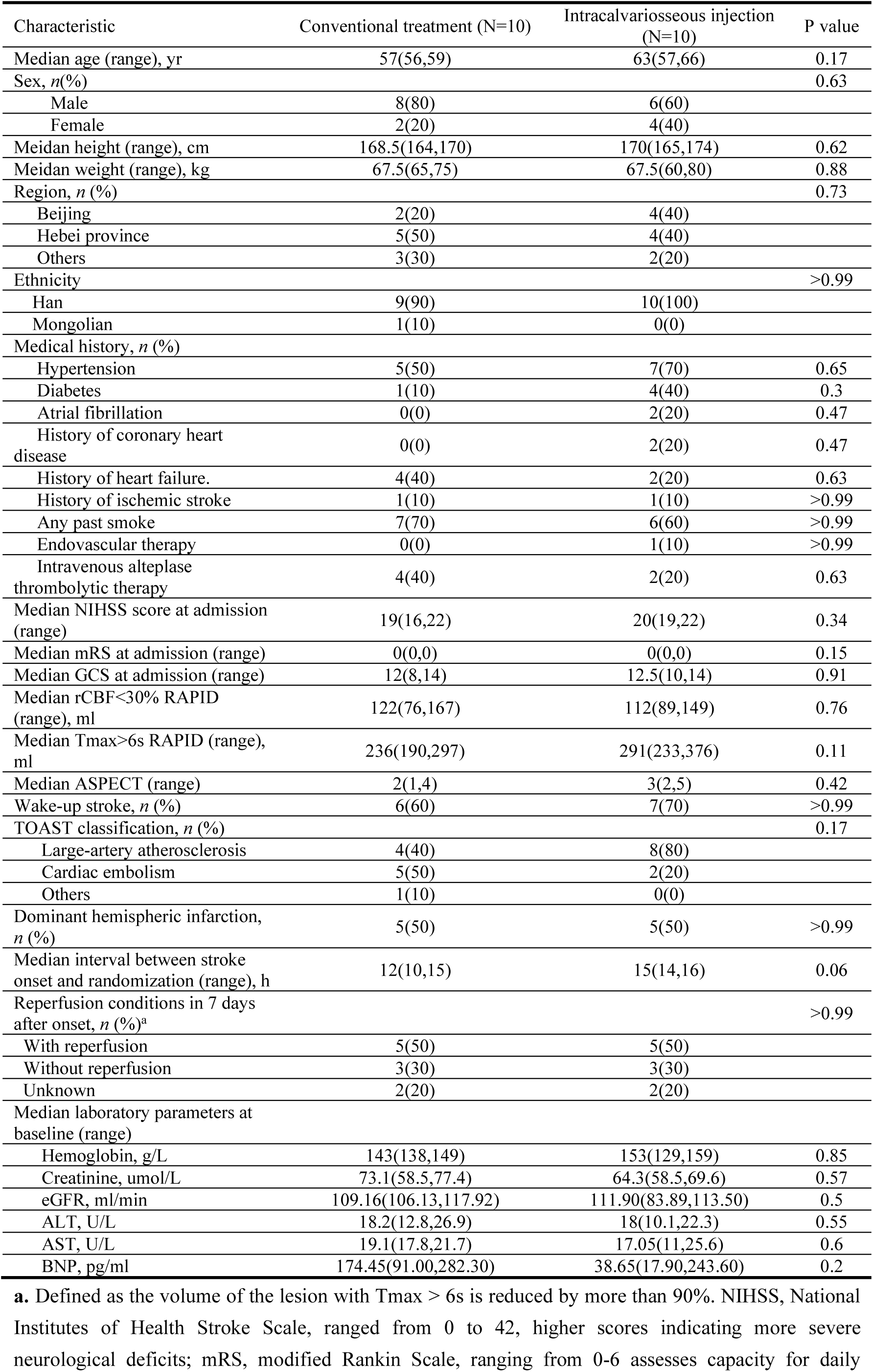
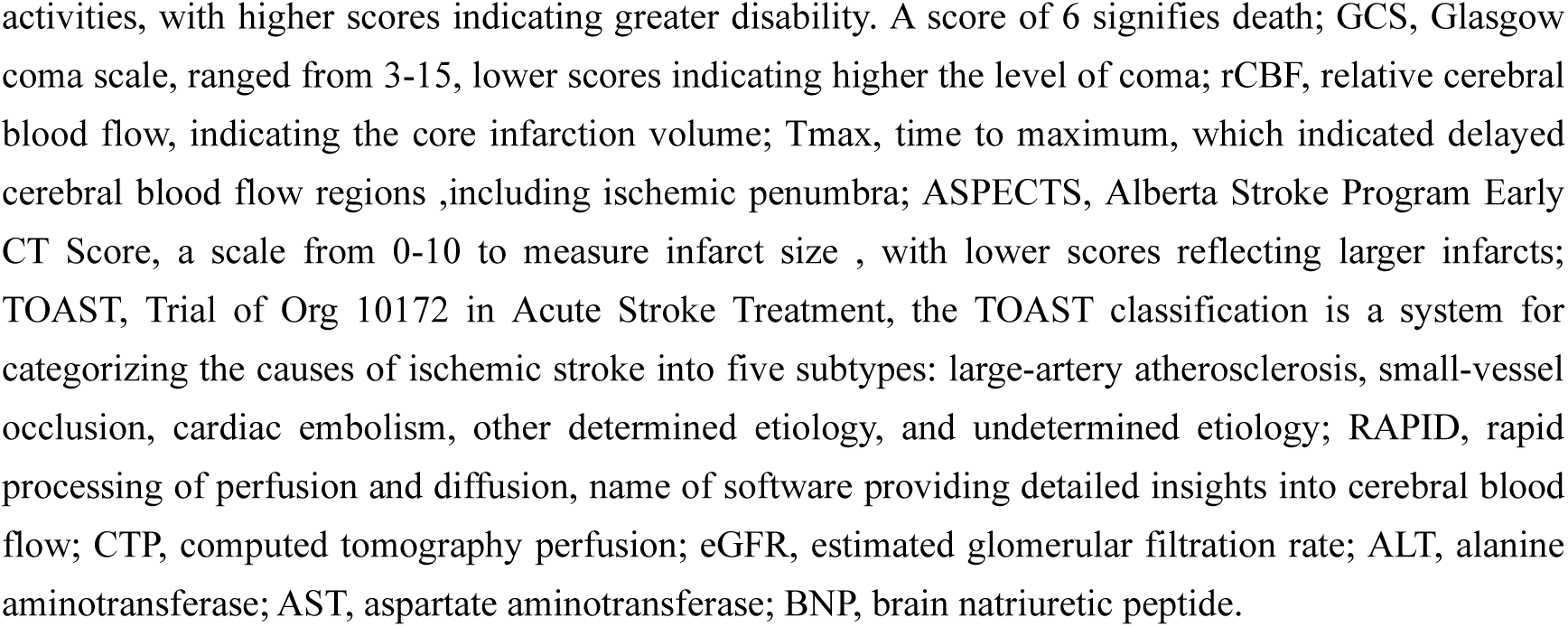
Clinical characteristics of the patients at baseline.

| Characteristic | Conventional treatment (N=10) | Intracalvariosseous injection (N=10) | P value |
| --- | --- | --- | --- |
| Median age (range), yr | 57(56,59) | 63(57,66) | 0.17 |
| Sex, *n*(%) |  |  | 0.63 |
| Male | 8(80) | 6(60) |  |
| Female | 2(20) | 4(40) |  |
| Meidan height (range), cm | 168.5(164,170) | 170(165,174) | 0.62 |
| Meidan weight (range), kg | 67.5(65,75) | 67.5(60,80) | 0.88 |
| Region, *n* (%) |  |  | 0.73 |
| Beijing | 2(20) | 4(40) |  |
| Hebei province | 5(50) | 4(40) |  |
| Others | 3(30) | 2(20) |  |
| Ethnicity |  |  | >0.99 |
| Han | 9(90) | 10(100) |  |
| Mongolian | 1(10) | 0(0) |  |
| Medical history, *n* (%) |  |  |  |
| Hypertension | 5(50) | 7(70) | 0.65 |
| Diabetes | 1(10) | 4(40) | 0.3 |
| Atrial fibrillation | 0(0) | 2(20) | 0.47 |
| History of coronary heart disease | 0(0) | 2(20) | 0.47 |
| History of heart failure. | 4(40) | 2(20) | 0.63 |
| History of ischemic stroke | 1(10) | 1(10) | >0.99 |
| Any past smoke | 7(70) | 6(60) | >0.99 |
| Endovascular therapy | 0(0) | 1(10) | >0.99 |
| Intravenous alteplase thrombolytic therapy | 4(40) | 2(20) | 0.63 |
| Median NIHSS score at admission (range) | 19(16,22) | 20(19,22) | 0.34 |
| Median mRS at admission (range) | 0(0,0) | 0(0,0) | 0.15 |
| Median GCS at admission (range) | 12(8,14) | 12.5(10,14) | 0.91 |
| Median rCBF<30% RAPID (range), ml | 122(76,167) | 112(89,149) | 0.76 |
| Median Tmax>6s RAPID (range), ml | 236(190,297) | 291(233,376) | 0.11 |
| Median ASPECT (range) | 2(1,4) | 3(2,5) | 0.42 |
| Wake-up stroke, *n* (%) | 6(60) | 7(70) | >0.99 |
| TOAST classification, *n* (%) |  |  | 0.17 |
| Large-artery atherosclerosis | 4(40) | 8(80) |  |
| Cardiac embolism | 5(50) | 2(20) |  |
| Others | 1(10) | 0(0) |  |
| Dominant hemispheric infarction, *n* (%) | 5(50) | 5(50) | >0.99 |
| Median interval between stroke onset and randomization (range), h | 12(10,15) | 15(14,16) | 0.06 |
| Reperfusion conditions in 7 days after onset, *n* (%)<sup>a</sup> |  |  | >0.99 |
| With reperfusion | 5(50) | 5(50) |  |
| Without reperfusion | 3(30) | 3(30) |  |
| Unknown | 2(20) | 2(20) |  |
| Median laboratory parameters at baseline (range) |  |  |  |
| Hemoglobin, g/L | 143(138,149) | 153(129,159) | 0.85 |
| Creatinine, umol/L | 73.1(58.5,77.4) | 64.3(58.5,69.6) | 0.57 |
| eGFR, ml/min | 109.16(106.13,117.92) | 111.90(83.89,113.50) | 0.5 |
| ALT, U/L | 18.2(12.8,26.9) | 18(10.1,22.3) | 0.55 |
| AST, U/L | 19.1(17.8,21.7) | 17.05(11,25.6) | 0.6 |
| BNP, pg/ml | 174.45(91.00,282.30) | 38.65(17.90,243.60) | 0.2 |

**a.** Defined as the volume of the lesion with Tmax > 6s is reduced by more than 90%. NIHSS, National Institutes of Health Stroke Scale, ranged from 0 to 42, higher scores indicating more severe neurological deficits; mRS, modified Rankin Scale, ranging from 0-6 assesses capacity for daily
activities, with higher scores indicating greater disability. A score of 6 signifies death; GCS, Glasgow coma scale, ranged from 3-15, lower scores indicating higher the level of coma; rCBF, relative cerebral blood flow, indicating the core infarction volume; Tmax, time to maximum, which indicated delayed cerebral blood flow regions ,including ischemic penumbra; ASPECTS, Alberta Stroke Program Early CT Score, a scale from 0-10 to measure infarct size , with lower scores reflecting larger infarcts; TOAST, Trial of Org 10172 in Acute Stroke Treatment, the TOAST classification is a system for categorizing the causes of ischemic stroke into five subtypes: large-artery atherosclerosis, small-vessel occlusion, cardiac embolism, other determined etiology, and undetermined etiology; RAPID, rapid processing of perfusion and diffusion, name of software providing detailed insights into cerebral blood flow; CTP, computed tomography perfusion; eGFR, estimated glomerular filtration rate; ALT, alanine aminotransferase; AST, aspartate aminotransferase; BNP, brain natriuretic peptide.

**Extended Data Table 2.**
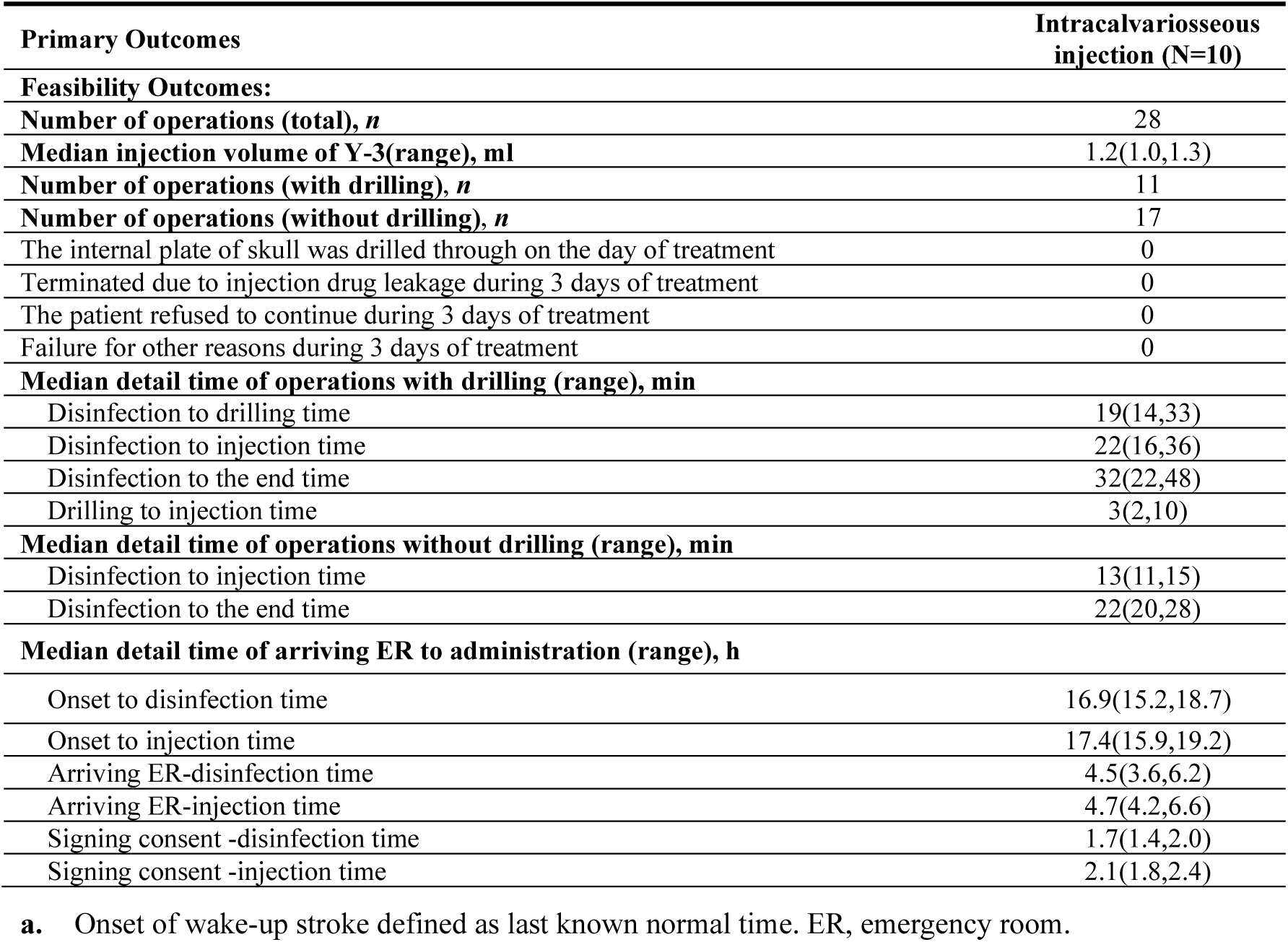
Feasibility outcomes and details of operations.

| **Primary Outcomes** | **Intracalvariosseous injection (N=10)** |
| --- | --- |
| **Feasibility Outcomes:** |  |
| **Number of operations (total), *n*** | 28 |
| **Median injection volume of Y-3(range), ml** | 1.2(1.0,1.3) |
| **Number of operations (with drilling), *n*** | 11 |
| **Number of operations (without drilling), *n*** | 17 |
| The internal plate of skull was drilled through on the day of treatment | 0 |
| Terminated due to injection drug leakage during 3 days of treatment | 0 |
| The patient refused to continue during 3 days of treatment | 0 |
| Failure for other reasons during 3 days of treatment | 0 |
| **Median detail time of operations with drilling (range), min** |  |
| Disinfection to drilling time | 19(14,33) |
| Disinfection to injection time | 22(16,36) |
| Disinfection to the end time | 32(22,48) |
| Drilling to injection time | 3(2,10) |
| **Median detail time of operations without drilling (range), min** |  |
| Disinfection to injection time | 13(11,15) |
| Disinfection to the end time | 22(20,28) |
| **Median detail time of arriving ER to administration (range), h** |  |
| Onset to disinfection time | 16.9(15.2,18.7) |
| Onset to injection time | 17.4(15.9,19.2) |
| Arriving ER-disinfection time | 4.5(3.6,6.2) |
| Arriving ER-injection time | 4.7(4.2,6.6) |
| Signing consent -disinfection time | 1.7(1.4,2.0) |
| Signing consent -injection time | 2.1(1.8,2.4) |

**a.** Onset of wake-up stroke defined as last known normal time. ER, emergency room.

**Extended Data Table 3.** Secondary efficacy outcomes.

| Secondary outcomes | Conventional treatment | Intracalvariosseous injection | Risk ratio/$\beta$ (95% CI) | P value |
| --- | --- | --- | --- | --- |
| efficacy outcomes (secondary): |  |  |  |  |
| Rate of patients with NIHSS improved 4 or more points from baseline at 7±2 days, *n* (%) | 1(10) | 2(20) | 2.00(0.21 to 18.69) | >0.99 |
| Rate of patients with NIHSS improved 4 or more points from baseline at 14±2 days, *n* (%) | 1(10) | 6(60) | 6.00 (0.87 to 41.21) | 0.057 |
| Rate of patients with NIHSS of limbs improved 2 or more points from baseline at 7±2 days, *n* (%) | 0(0) | 1(10) | NA | >0.99 |
| Rate of patients with NIHSS of limbs improved 2 or more points from baseline at 14±2 days, *n* (%) | 0(0) | 0(0) | NA | NA |
| Median change of rCBF<30% from baseline to 7±2 days in RAPID (IQR)—ml<sup>a</sup> | -61.5(-129.5, -30) | -91.5(-112,10) | 32.63(-97.75,163.00) | 0.60 |
| Median change of Tmax>6s from baseline to 7±2 days in RAPID (range), ml<sup>a</sup> | -116.5(-202.5,-75) | -190.5(-271.5,-21) | -23.88(-166.52,118.77) | 0.72 |
| Decompressive craniectomy, *n* (%) | 4(40) | 2(20) | 0.50(0.12,2.14) | 0.63 |
| early-discharge case, *n* (%)<sup>a</sup> | 2(20) | 2(20) | 1(0.17,5.78) | >0.99 |
| Median NICU hospitalization days (range)<sup>a</sup> | 12(6,19) | 14(8,16) | -1.20(-6.69,4.29) | 0.65 |
| Median cost of the NICU hospitalization (range), thousand yuan<sup>a</sup> | 72.33(34.0,102.5) | 56.48(33.5,88.3) | -7.84(-39.40,23.72) | 0.61 |

**a.** Conventional treatment group had two early-discharge cases, one of which died and the other survived. ICO group had two early-discharge cases which were both mortality cases. All these cases did not complete the 7-day CTP and were treated as missing values, but were included in the analysis of the NICU hospitalization days and costs. CT, Conventional treatment; ICO, Intracalvariosseous Injection; NIHSS, National Institutes of Health Stroke Scale, ranged from 0 to 42, higher scores indicating more severe neurological deficits; rCBF, relative cerebral blood flow; RAPID, rapid processing of perfusion and diffusion, name of software providing detailed insights into cerebral blood flow; GCS, Glasgow coma scale, ranged from 3-15, lower scores indicating higher the level of coma; CTP, computed tomography perfusion; NICU, Neuro intensive Care Unit; LOCF, last observation carried forward.

